# Behavior-driven forecasts of neighborhood-level COVID-19 spread in New York City

**DOI:** 10.1101/2024.04.17.24305995

**Authors:** Renquan Zhang, Jilei Tai, Qing Yao, Wan Yang, Kai Ruggeri, Jeffrey Shaman, Sen Pei

**Affiliations:** School of Mathematical Sciences, Dalian University of Technology, Dalian, 116024, China; Department of Environmental Health Sciences, Mailman School of Public Health, Columbia University, New York, NY 10032; Department of Epidemiology, Mailman School of Public Health, Columbia University, New York, NY 10032; Herbert Irving Comprehensive Cancer Center, Columbia University Medical Center, New York, NY 10032; Department of Health Policy and Management, Mailman School of Public Health, Columbia University, New York, NY 10032; Columbia Climate School, Columbia University, New York, NY 10027

## Abstract

The COVID-19 pandemic in New York City (NYC) was characterized by marked disparities in disease burdens across neighborhoods. Accurate neighborhood-level forecasts are critical for planning more equitable resource allocation to reduce health inequalities; however, such spatially high-resolution forecasts remain scarce in operational use. In this study, we analyze aggregated foot traffic data derived from mobile devices to measure the connectivity among 42 NYC neighborhoods driven by various human activities such as dining, shopping, and entertainment. Using real-world time-varying contact patterns in different place categories, we develop a parsimonious behavior-driven epidemic model that incorporates population mixing, indoor crowdedness, dwell time, and seasonality of virus transmissibility. We fit this model to neighborhood-level COVID-19 case data in NYC and further couple this model with a data assimilation algorithm to generate short-term forecasts of neighborhood-level COVID-19 cases in 2020. We find differential contact patterns and connectivity between neighborhoods driven by different human activities. The behavior-driven model supports accurate modeling of neighborhood-level SARS-CoV-2 transmission throughout 2020. In the best-fitting model, we estimate that the force of infection (FOI) in indoor settings increases sublinearly with crowdedness and dwell time. Retrospective forecasting demonstrates that this behavior-driven model generates improved short-term forecasts in NYC neighborhoods compared to several baseline models. Our findings indicate that aggregated foot-traffic data for routine human activities can support neighborhood-level COVID-19 forecasts in NYC. This behavior-driven model may be adapted for use with other respiratory pathogens sharing similar transmission routes.

**Author summary:** A fundamental question in infectious disease modeling is whether the inclusion of more detailed processes results in more precise epidemic simulation, and to what extent system granularity is needed to inform real-world application of model outcomes. Here, we investigate the utility of foot traffic data, which capture mobility patterns during various human activities, in neighborhood-level disease modeling. Our results indicate that foot traffic data aggregated for place categories, combined with representation of the seasonality of SARS-CoV-2 transmissibility, can support forecasting of the heterogeneous COVID-19 spread across 42 NYC neighborhoods in 2020. We propose a parsimonious behavior-driven epidemic model that generates improved short-term forecasts at the neighborhood level. We also estimate that the force of infection (FOI) in indoor settings increases sublinearly with crowdedness and dwell time. Incorporating the differential FOIs in different place categories improves short-term COVID-19 forecasts. With proper modifications, this model could be applied to other respiratory diseases.

## Introduction

In urban settings, COVID-19 has imposed differential disease and healthcare burdens across neighborhoods characterized by varying demographic and socioeconomic conditions [1–3]. A case in point is New York City (NYC), where pronounced health disparities within the city have been reported [4–7]. The stark difference in disease burdens requires the delivery of tailored services and support to local communities. Accurate forecasting of respiratory disease outbreaks at the neighborhood level can inform policymaking to design more equitable interventions and resource allocation in future epidemics. Nonetheless, generating such spatially high-resolution forecasts remain challenging due to the significant behavioral variability and interconnectedness among neighborhoods in metropolitan areas.

The transmission dynamics of SARS-CoV-2 were collectively shaped by human behavior, seasonal variation in virus transmissibility (i.e., seasonality), population immunity, and pathogen evolution [8]. Among those factors, human behaviors, both voluntary and those driven by governmental interventions, played a major role during the early phase of the pandemic [9–15]. Many studies used mobility data to produce short-term COVID-19 forecasts and some attempted to determine the added value of including mobility data above a baseline model [16–19]. Despite these efforts, a key question remains – what metrics of mobility should be used in epidemic modeling? Early studies examining the associations between human mobility indices (e.g., distance traveled, time spent at home, or percentage change in mobility) and COVID-19 spread yielded inconclusive results – strong associations during lockdowns [20–22] but weak or no associations during the summer of 2020 [23,24]. A potential explanation for this inconsistency is that the mobility indices employed did not accurately reflect behavioral effects on the contagion process. For instance, a person can drive a long distance alone without contributing to pathogen transmission, yet a short visit to a convenience store may involve multiple exposures. As a result, incorporating mobility indices into process-based models may not necessarily improve disease forecasting.

Foot traffic data derived from mobile devices offer an effective means of tracking human mobility and behavior patterns. Unlike aggregated mobility indices, foot traffic data document visitation records at points of interest (POIs) with high spatial and temporal resolution, providing a direct measurement of population mixing in various locations. These privacy-preserving data have supported studies on high-resolution disease modeling [25–29], risk of airborne transmission [30], and human mobility changes during outbreaks or natural disasters [31–33]. For instance, mobility network models and agent-based models were used to simulate epidemic spread within small geographical units, which provided valuable insights on inequalities of infection risk across communities [25–29]. Another study used foot-traffic data to model the transmission of SARS-CoV-2 at the sub-county scale [26]. However, it remains unclear whether these models can reproduce heterogeneous SARS-CoV-2 transmission at the neighborhood level in urban settings and support forecasting with validated skill. In addition, the complexity of many highly detailed models makes it challenging for rapid deployment by public health officials during an emergency.

In this study, we aim to develop a parsimonious process-based model informed by aggregated foot traffic data to generate validated neighborhood-level forecasts with efficient simulation and calibration. We analyzed foot traffic data within various place categories in NYC during 2020. Densely populated metropolitan areas like NYC often experience earlier outbreaks, which may then propagate rapidly to other locations. During the COVID-19 pandemic, NYC was the first epicenter in the US. Using fine-grained foot traffic data, we identified distinct mobility and indoor contact patterns across five place categories: restaurants and bars, retail, arts and entertainment, educational settings, and other places. We then developed a behavior-driven epidemic model informed by category-specific mobility and indoor contact pattern data. Our results indicate that this model, capturing social drivers of contagion (i.e., human contacts in various activities) and transmission seasonality, can fit COVID-19 spread across the 42 NYC neighborhoods during 2020. Furthermore, we demonstrated that this model, in conjunction with data assimilation techniques, produced improved short-term forecasts of COVID-19 spread at the neighborhood level relative to three baseline models with simpler constructs, considering four metrics - mean absolute percentage error, log score, weighted interval score, and the coverage of 95% predictive intervals.

## Results

### Mobility and contact patterns driven by human activities

Human contacts are driven by various activities such as dining, shopping, work, and entertainment. We used foot traffic data shared by SafeGraph [34] to track mobility and contact patterns during these activities (Materials and Methods). Using the North American Industry Classification System (NAICS) code [35], we classified all POIs into five categories: (1) restaurants and bars, (2) retail, (3) arts and entertainment, (4) educational settings, and (5) other places (Table S1). A clustering analysis indicates that POIs within each category exhibit similar crowdedness, dwell time, and variance of visitor numbers during a week (Fig. S1). This categorical classification allows parsimonious representation of population mixing in different settings and aligns with the practical implementation of governmental interventions targeting specific high-risk sectors.

The foot traffic data recorded the overall temporal trend of mobility in NYC. The daily number of visitors to all place categories plummeted in March 2020 and was reduced by 81% at its lowest point on April 12, 2020 (Fig. 1A), following strict control measures enforced during the initial wave (see the daily reported COVID-19 cases in Fig. 1B). Thereafter, mobility gradually increased throughout 2020 but did not recover to its pre-pandemic level.

**Fig. 1.**
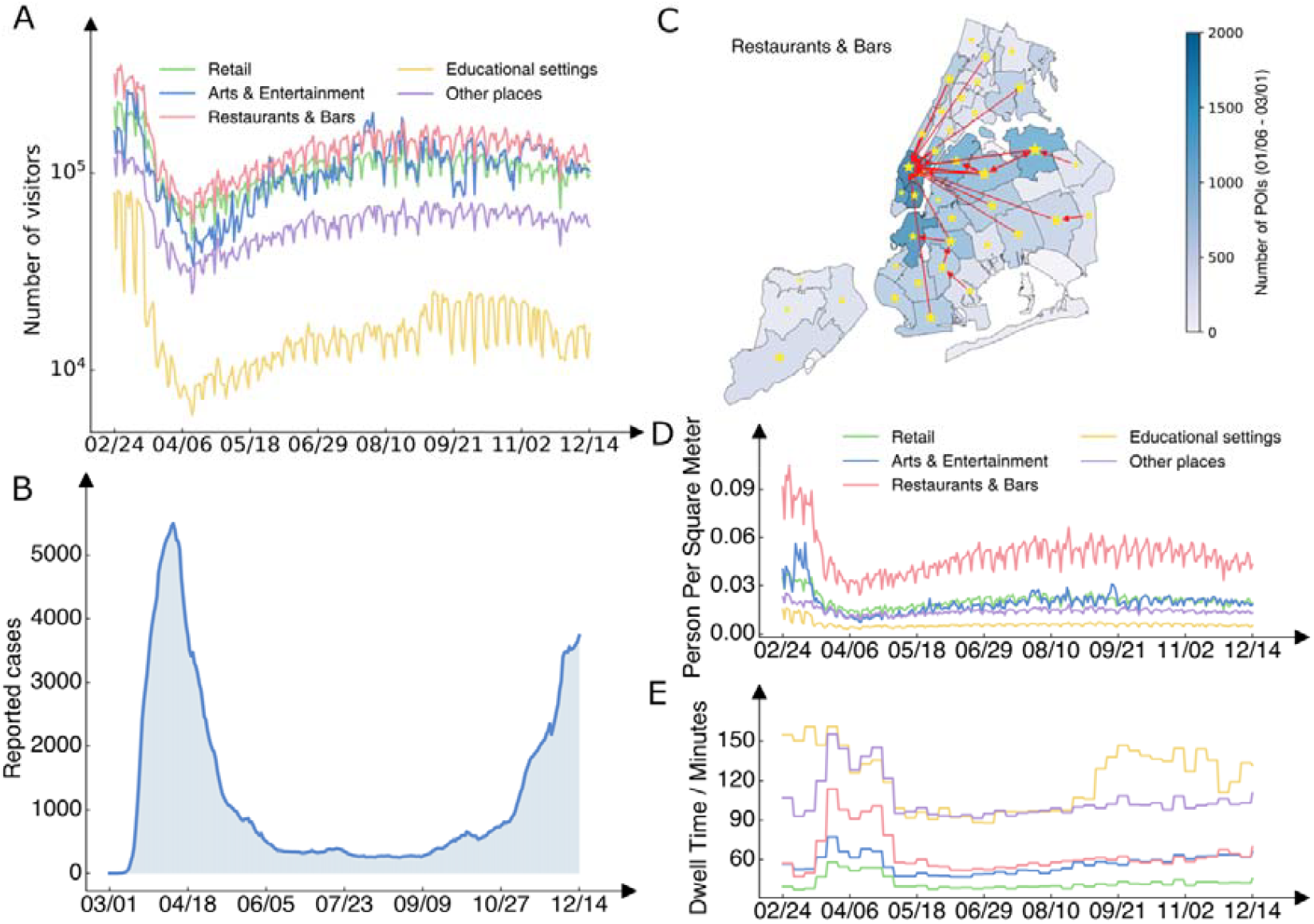
Mobility and contact patterns in different place categories. (A), Daily visitor counts to five place categories (restaurants & bars, retail, arts & entertainment, educational settings, and others) in NYC during 2020, as recorded in the foot traffic data. (B), Daily reported COVID-19 cases in NYC. (C), Geographical distribution of restaurants & bars across NYC neighborhoods (color). Stars and arrows highlight mobility links with over 1000 visitors per day from January 6, 2020 to March 1, 2020. Stars indicate self-links where residents visited restaurants & bars within their own neighborhoods. The maps were created using PYTHON using the shapefile publicly available at https://github.com/nychealth/coronavirus-data/tree/master/Geography-resources. This is a public repository by NYC Department of Health and Mental Hygiene. The term of use can be found here: https://github.com/nychealth/coronavirus-data. (D), Daily average crowdedness (daily visitor counts per square meter) at POIs for five place categories. (E), Daily average dwell time (minutes) at POIs for five place categories.

The foot traffic data provided insights on the distribution of built environments within NYC. The NYC Department of Health and Mental Hygiene (DOHMH) uses United Hospital Fund (UHF) Areas as one way to designate neighborhoods (Fig. 1C). As the spatial allocation of POIs can significantly influence the mobility of residents engaged in various activities, we examined the number of POIs located within each neighborhood. We observed heterogeneous geographical distributions of POIs and distinct spatial patterns across place categories (Fig. S2), revealing spatial disparities in urban services and facilities. For instance, restaurants and bars concentrated in lower Manhattan and drew visitors from distant neighborhoods (Fig. 1C), whereas educational facilities exhibited a more even distribution across neighborhoods (Fig. S2).

The foot traffic data captured the interconnectedness between neighborhoods driven by different human activities. To assess population mixing between the 42 UHF neighborhoods, we constructed daily mobility matrices for each place category, denoted by 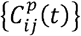, where 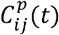 represents the number of visitors from home neighborhood *i* to destination neighborhood *j* in place category *p* on day *t*. We visualized average daily mobility for four place categories (excluding other places that lump in different POI types) from January 6, 2020, to March 1, 2020 (Fig. 2), depicting mobility patterns in winter prior to the pandemic. Notably, regardless of place categories, most visitors went to POIs in their home neighborhoods, as evidenced by the diagonal elements with large daily visitor counts. A clear cross-neighborhood mobility structure emerged within four boroughs (the Bronx, Brooklyn, Queens, and Staten Island), indicating that residents in these boroughs frequently visited local POIs close to their residence. In contrast, Manhattan, the primary employment borough with high living costs [36], drew large numbers of visitors from other boroughs.

**Fig. 2.**
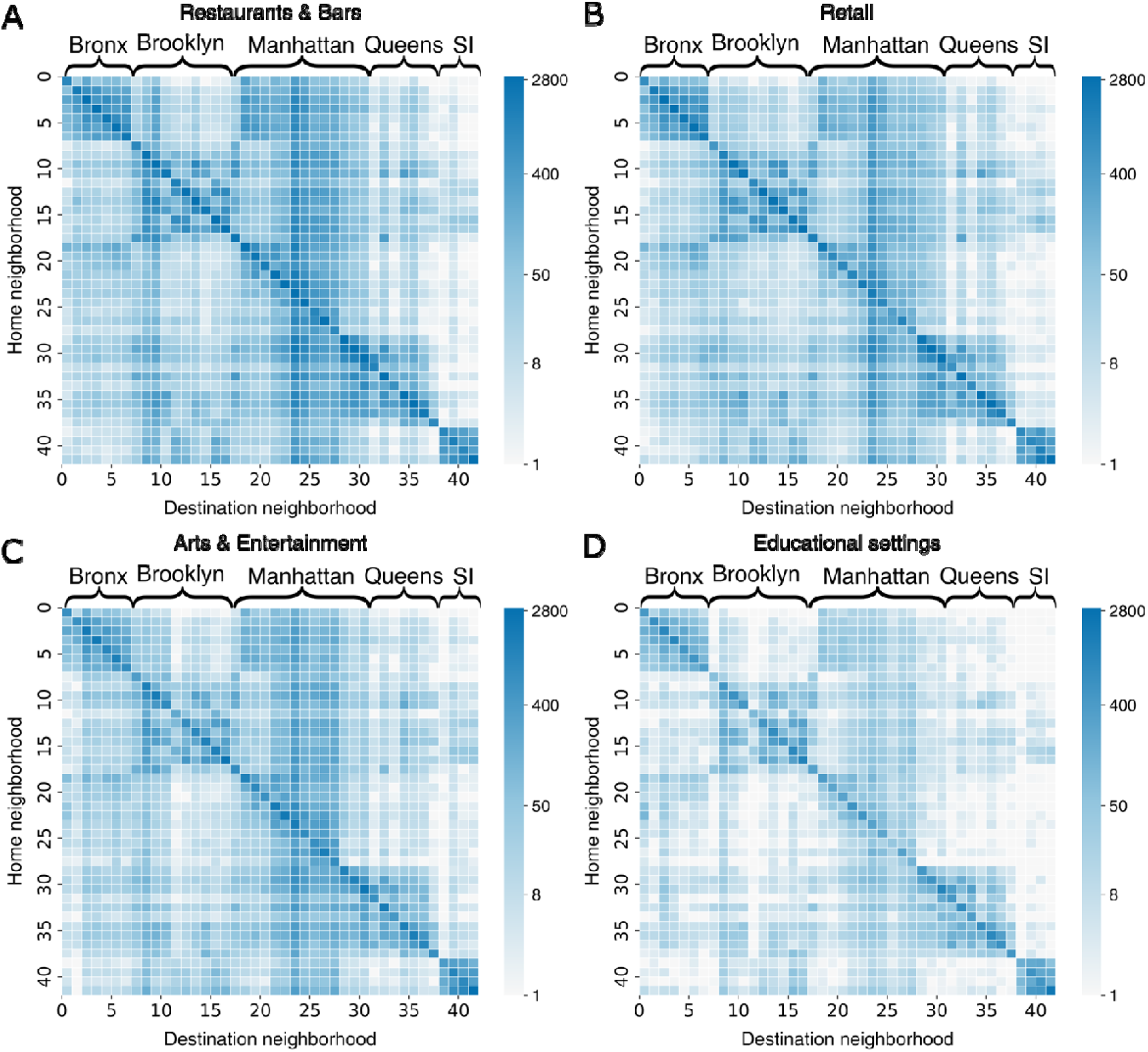
Mobility patterns across 42 NYC neighborhoods in four place categories. Daily average visitor counts (in log scale) from home neighborhoods (y-axis) to destination neighborhoods (x-axis) in restaurants & bars (A), retail (B), arts & entertainment (C), and educational settings (D). The five boroughs of NYC (the Bronx, Brooklyn, Manhattan, Queens, and Staten Island) are indicated on top of each heatmap. Foot traffic data from January 6, 2020 to March 1, 2020 were used, representing the period before the implementation of governmental interventions. To avoid numerical issues for, we visualized the quantity, where is the daily average visitor count.

Cross-neighborhood mobility was most pronounced for restaurants and bars and least notable for educational facilities. We compiled daily mobility matrices for 2020 to track the temporal evolution of cross-neighborhood population mixing.

The foot traffic data also revealed contact patterns within POIs that may modulate infection risk in indoor environments. Particularly, we focused on the crowdedness (measured by daily number of visitors per square meter) [37] and average dwell time of visitors (Materials and Methods) [25]. Among the five examined place categories, restaurants and bars had on average the highest crowdedness (Fig. 1D). The crowdedness in all place categories decreased by up to 82% (arts and entertainment) following the lockdown in March 2020 and then moderately increased in the latter half of 2020. In contrast, dwell time in all place categories showed less variation (Fig. 1E). The increase in average dwell time during the lockdown for most place categories seems counterintuitive; however, those longer dwell time are for the smaller number of people who remained present in those settings, such as employees. We also found that the average dwell time in educational facilities increased starting in September, corresponding to the start of the fall school semester. Variation in crowdedness and dwell time may lead to differential infection risk in POIs.

### A behavior-driven epidemic model

To model contagion processes driven by human activities, we developed a behavior-driven epidemic model informed by real-world mobility matrices for different place categories, time-varying crowdedness and dwell time, and seasonality of virus transmissibility. Denote 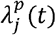 as the force of infection (FOI) in place category *p* in neighborhood *j* on day *t*. The transmission dynamics are described by the following equations.

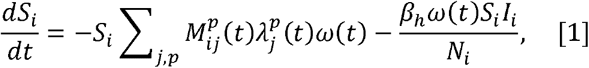

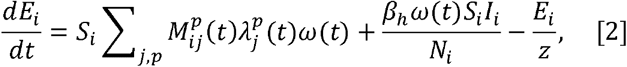

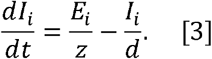

Here *N*_*i*_, *s*_*i*_, *t*_*i*_, and *I*_*i*_ are the total, susceptible, exposed, and infectious population in neighborhood *i*; *z* and *d* are the latency and infectious duration; *β*_*h*_ is the baseline transmission rate in places not captured by the foot traffic data; 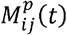 represents the fraction of population living in neighborhood *i* visiting place category *p* in neighborhood *j* on day *t*; 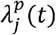 is the force of infection (FOI) in place category *p* in neighborhood *j* on day *t*, parameterized using crowdedness and dwell time; and *ω*(*t*) imposes seasonality on the transmissibility of SARS-CoV-2.

We linked FOI to crowdedness and dwell time through:

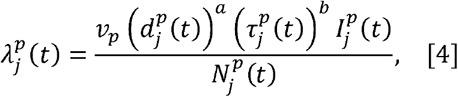

where 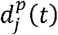 and 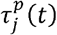 are the crowdedness and average dwell time in place category *p* in neighborhood *j* on day. 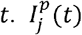 and 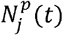 denote the infectious and total population present in place category *p* in neighborhood *j* on day *t*. Both 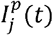 and 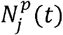 are computed using the mobility matrices 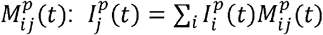 and 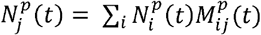. The parameters *a* and *b* determine the nonlinear relationship between virus transmissibility and crowdedness and dwell time, respectively. Note, *a* and *b* are shared by all place categories, representing the universal functional form linking crowdedness and dwell time to FOI that applies in all POIs. In practice, this functional form links real-world intervention criteria such as occupancy limit and operation hours, which are straightforward to measure and control, to the transmission risk of infectious diseases. We further introduced a multiplier, *v*_*p*_, for each place category representing the compound effects of potential sampling bias across different place categories, category-specific interventions, relative contribution to infection risk in different place categories, and other unaccounted factors. Note that an individual may be counted as a visitor to multiple place categories on the same day; however, the relative infection risk in multiple place categories on the same day (e.g., modulated by the time spent in different place categories) can be factored in the parameter *v*_*p*_ and estimated through model fitting. As a result, this model structure addresses the issue of double-counting susceptible population in each neighborhood.

In addition to population mixing, the transmissibility of SARS-CoV-2 exhibits strong seasonality modulated by meteorological factors, with humidity playing the major role in the northern US [38]. We imposed a seasonality term, *ω*(*t*), forced by absolute humidity (AH), derived from the modeling of influenza seasonality [39,40]. Specifically, we define *ω*(*t*) ∝ *w*_*min*_ + exp (−180 × *q*(*t*) log (*w*_*max*_ – *w*_*min*_)), where *w*_*max*_ and *w*_*min*_ are the parameters controlling the intensity of seasonality, and *q*(*t*) is the time-varying specific humidity, a measure of absolute humidity. This functional form has been validated for both influenza modeling and forecasting [39,40] (see a detailed discussion on seasonality in Materials and Methods). We normalized *ω* (*t*)so that the average value over a year is one. For the first year of the pandemic, we did not consider immunity loss as studies showed re-infections were rare [41]. We further modeled the time-varying ascertainment rate [42] (Fig. S3) and delay from infection to case confirmation (Materials and Methods). The model was integrated daily using the Euler stepping scheme deterministically.

### Estimating the dependency of FOIs on crowdedness and dwell time

We first used grid search and model selection to estimate the parameters *a* and *b* that control the impacts of crowdedness and dwell time on FOIs. We tested 31 × 31 parameter combinations: *a* ∈,[0,3] and *b* ∈,[0,3] with a step of 0.1 for both parameters. For each parameter combination, we fixed *a* and *b* in the model and calibrated the model against real-world weekly COVID-19 cases in 42 neighborhoods from March 1, 2020 to June 7, 2020, a period with substantial variation in crowdedness and dwell time (Fig. 1). Model calibration was performed using Metropolis-Hastings Markov Chain Monte Carlo (MCMC) (Materials and Methods) [43]. The goodness-of-fit for each parameter combination was measured using log-likelihood (Materials and Methods). The landscape of log-likelihood indicates that the best-fitting model was roughly in the region of *a* ∈,[0,1] and *b* ∈,[0,1] (Fig. S4).

To more accurately estimate the parameters *a* and *b*, we used a Bayesian approach to obtain the uncertainty of these parameters (see technical details in Supplementary Information). The median posterior estimates for the parameters *a* and *b* are *a* = 0.30 (95% CI [0.19,0.403]) and *b* = 0.13 (95% CI [0.03, 0.22]). The median estimates suggest that FOIs likely increases sublinearly with crowdedness and dwell time. In other words, for the median estimates, pathogen transmission risk will increase rapidly when crowdedness and dwell time rise from lower values; however, the rate of increase declines for greater crowdedness and longer dwell time, showing a diminishing-return effect. In the following MCMC model fitting to the COVID-19 data in 2020, we fixed *a* and *b* at the median estimates; however, in retrospective forecasting, we randomly drew parameters *a* and *b* from the 95% posterior CIs of the ensemble members, ensuring that model predictions captured the uncertainty in these two parameters. Note, it is preferable to sample *a* and *b* from the joint posterior distribution. However, as *a* and *b* do not have a strong correlation in the posterior, we do not expect substantial changes in forecasting performance.

### Modeling neighborhood-level COVID-19 spread

We fit the behavior-driven epidemic model to weekly COVID-19 cases in 42 neighborhoods from March 1, 2020 to December 13, 2020, before the availability of COVID-19 vaccines in NYC. For model fitting, we fixed epidemiological and seasonality parameters and estimated *v*_*p*_ for five place categories, *β*_*h*_ for baseline transmission, and a parameter *h* controlling the magnitude of observation error. Model fitting was performed for the period from March 1, 2020 to December 13, 2020 using Metropolis-Hastings MCMC. Trace plots, the Gelman-Rubin 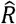 statistic, and effective sample size were used to assess the convergence of MCMC chains (Materials and Methods, Fig. S5). We also performed multiple chains starting from randomly drawn initial infections and found the results remained similar. The estimated posterior distributions for all parameters are shown in Fig. S6. To check the model goodness-of-fitting, we ran independent model simulations using samples from estimated posterior parameters and compared the simulated COVID-19 case counts with reported numbers from the 42 neighborhoods.

At the city level, the behavior-driven epidemic model generally reproduced the trend of COVID-19 cases throughout 2020 (Fig. 3A). In addition, model simulations agreed with observations in most neighborhoods (Fig. S7), suggesting that aggregated foot traffic data combined with seasonality can support neighborhood-level modeling of SARS-CoV-2 transmission. Model fitting in several neighborhoods such as Greenwich Village-SoHo had larger discrepancy, possibly due to model misspecification or issues in the mobility data.

**Fig. 3.**
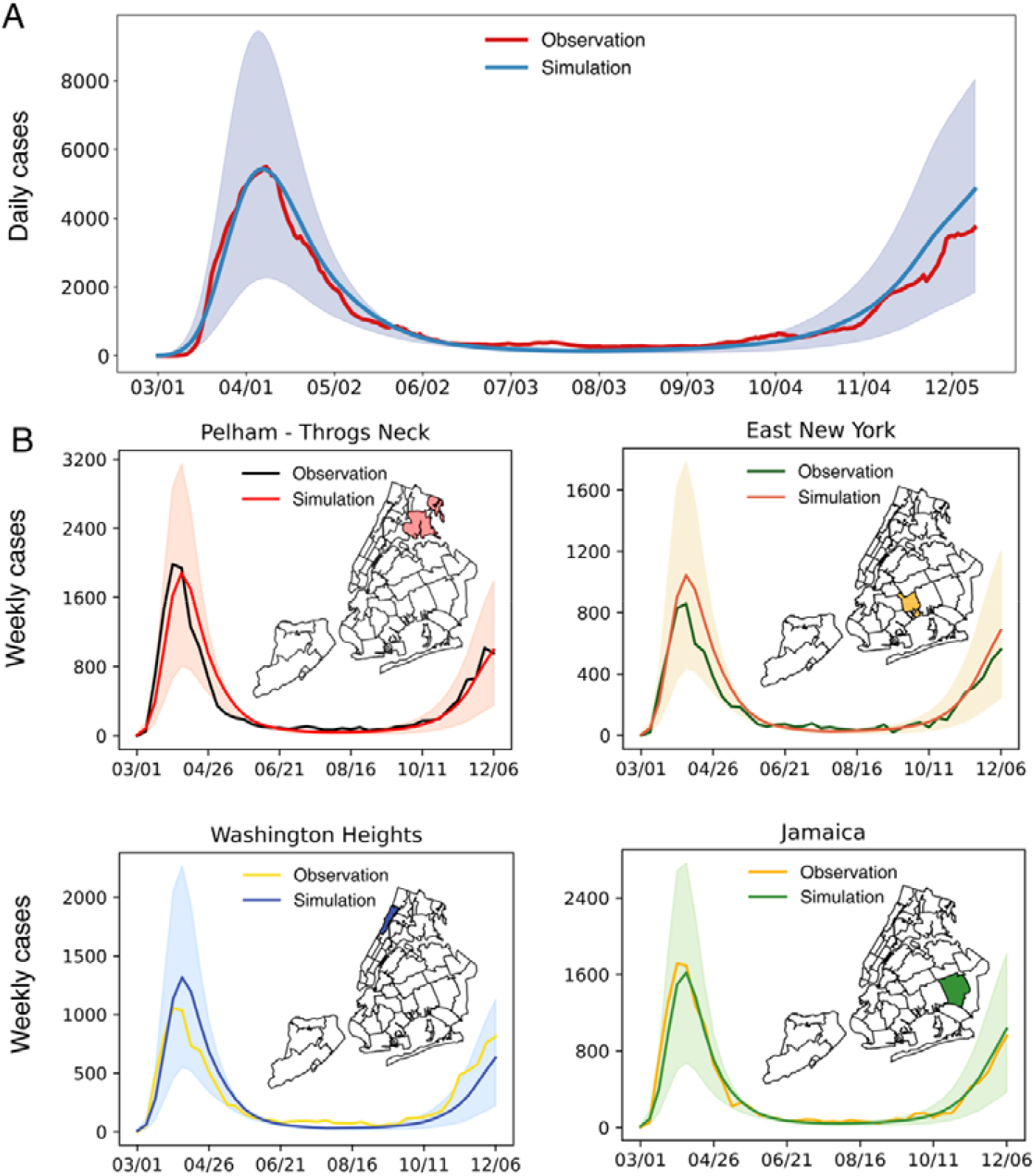
Model fitting to neighborhood-level COVID-19 case data. (A), Simulations using model parameters estimated for the period from March 1, 2020 to December 13, 2020. Simulated cases were aggregated to the city level and are compared with the daily confirmed cases in NYC (red line). The shaded blue area represents the 95% credible interval, obtained from 500 independent simulations, without adding the Gaussian observation error (i.e., it only reflects the uncertainty in the latent deterministic trajectories). The solid lines represent the median of those 500 trajectories. (B), Simulations in four representative neighborhoods in the Bronx (upper left), Brooklyn (upper right), Manhattan (lower left), and Queens (lower right). Maps display the geographical locations of these neighborhoods. The maps were created using PYTHON using the shapefile publicly available at https://github.com/nychealth/coronavirus-data/tree/master/Geography-resources. This is a public repository by NYC Department of Health and Mental Hygiene. The term of use can be found here: https://github.com/nychealth/coronavirus-data.

Examples in four neighborhoods in the Bronx, Brooklyn, Manhattan, and Queens are provided in Fig. 3B. To better visualize model fitting for cases spanning several orders of magnitude, we show the fitting performance in the log scale of the COVID-19 cases (Fig. S8), as suggested in [44]. Using model simulations in Fig. 3, we estimated that restaurants and bars and the baseline transmission contributed 76% and 17% of infections during the study period. The dominant role of restaurants and bars in COVID-19 spread was noted in previous works [25]. For our model, the large proportion of infections linked to restaurants and bars was likely caused by the dominating visiting records to this category in the foot-traffic data and the parametric flexibility to find the best proxies for transmission in mobility data.

To assess the impact of crowdedness and dwell time on transmission dynamics, we performed another fitting fixing *a* = 0 and *b* = 0 in the behavior-driven model. Without the dynamical constraint imposed by crowdedness and dwell time data, the model became more flexible. While this simpler model performed well for the spring wave, it significantly overestimated COVID-19 cases in the winter of 2020 (Figs. S9-S10). We speculate that this might be because the model with *a* =*b* = 0 did not capture the reduction of crowdedness in indoor settings following the policy of limiting capacity in certain businesses.

### Retrospective forecasts of neighborhood-level COVID-19 cases

While the behavior-driven epidemic model can reproduce real-world neighborhood-level SARS-CoV-2 transmission, it is unknown if this model can generate more accurate short-term forecasts of COVID-19 cases. We coupled the behavior-driven epidemic model with an efficient data assimilation algorithm, the ensemble adjustment Kalman filter (EAKF) [45], to generate retrospective forecasts at the neighborhood level (Materials and Methods, Supplementary Information). During the forecasting process, model parameters and variables were updated weekly once real-world COVID-19 surveillance data became available. We then integrated the optimized model into the future to generate forecasts. Within the forecasting horizon, mobility matrices, indoor contact patterns, and reporting rates were fixed using the information available when the forecasts were generated. We started retrospective forecasting on June 8, 2020, assuming we have the early-stage data to estimate key epidemiological parameters.

The one-week ahead forecasts of COVID-19 cases generally agreed with observations at the neighborhood level (Fig. S11), capturing the large variations in disease burden. We compared the forecast skill of the behavior-driven epidemic model with three baseline models: (B1), a metapopulation model without place category-specific mobility but with seasonal forcing; (B2), the behavior-driven model without seasonal forcing; (B3), the behavior driven model with seasonal forcing but static mobility matrices, crowdedness, and dwell time, averaged over the week prior to June 8, 2020 (before the start of forecasts) (Materials and Methods). The baseline models used the same model-EAKF forecasting framework and mobility information from the same foot traffic data to generate predictions. A similar framework has been used to model COVID-19 spread at the US county level [46,47]. The baseline models had decent forecasting performance at the neighborhood level (Figs. S12-S14).

We compared the one-week ahead forecasts generated for each neighborhood at each week using four metrics: (1) mean absolute percentage error (MAPE) for point predictions, (2) log score and (3) weighted interval score (WIS) [48] for probabilistic predictions (Materials and Methods), and (4) coverage for 95% predictive intervals. Log score and WIS measure the forecasting performance of probabilistic predictions, taking into account the uncertainty of predictive distributions. They have been used as the standard metrics to evaluate real-time forecasts for influenza [49] and COVID-19 [50]. Specifically, lower MAPEs, higher log scores, and lower WIS scores indicate better forecasts. In general, the proposed model generated more accurate forecasts in most neighborhoods and forecast weeks compared to B1 and B2, but not consistently (Fig. 4). When compared with B3, the proposed model performed better in the winter of 2020 but worked less well in the summer of 2020. The coverage of the behavior-driven model forecasts was comparable to or slightly better than those from the baseline models. The coverage for the 95% predictive intervals was 0.891 for the proposed model; for the baseline models, they were 0.864 for B1, 0.875 for B2, and 0.898 for B3. All models were overconfident at the 95% predictive intervals. Post-processing the spread of predictive intervals (e.g., inflating or deflating predictive intervals based on historical coverage performance) may improve the calibration of forecasts, as we have done in a previous work [51]. In certain periods and neighborhoods, simpler models had better performance than the behavior-driven model, particularly for B3. Furthermore, different metrics favored different models for a sizeable periods and neighborhoods. There was no single model that performed consistently best across all forecast weeks and neighborhoods. This finding points to the potential benefits from ensembling different models, in line with findings from previous forecasting initiatives [50]. We further compared two-week ahead forecasts (Fig. S15) and found that the findings remained similar.

**Fig. 4.**
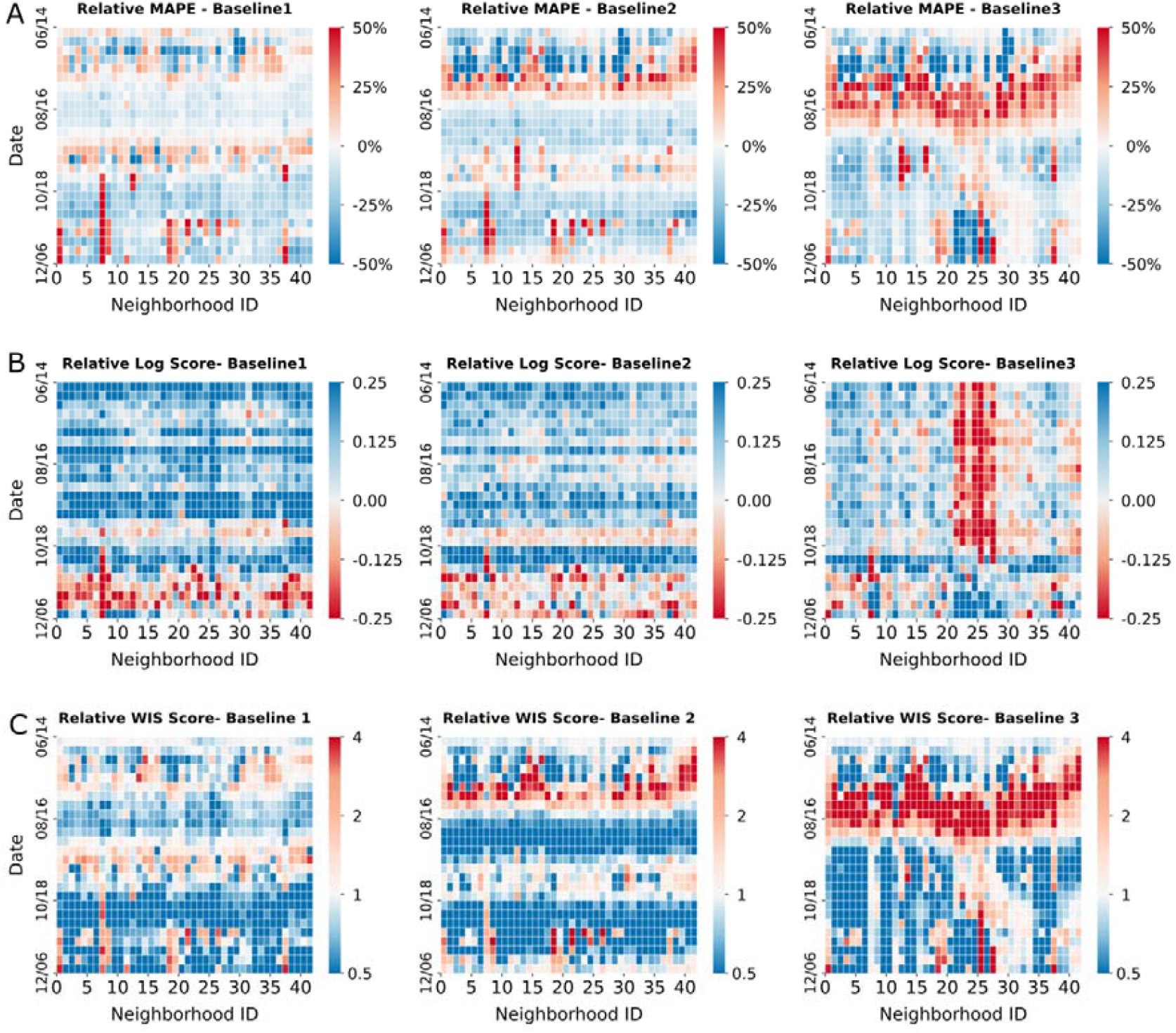
One-week ahead retrospective forecasts for neighborhood-level COVID-19 cases. The behavior-driven forecasts are compared with three baseline models: (B1), a metapopulation model without place category-specific mobility but with seasonal forcing; (B2), the behavior-driven model without seasonal forcing; (B3), the behavior driven model with seasonal forcing but static mobility matrices, crowdedness, and dwell time. We present (A) the relative mean absolute percentage error (MAPE) (MAPEs of the behavior-driven model minus those of the baselines, with blue indicating better forecasts), (B) relative log score (log scores of the behavior-driven model minus those of the baselines, with blue indicating better forecasts), and (C) relative weighted interval score (WIS) (the *ratio* of the WIS scores of the behavior-driven model to those of the baselines, with blue indicating better forecasts) for all 42 neighborhoods from June 8, 2020 to December 13, 2020.

## Discussion

Population contacts driven by human activities can facilitate the transmission of SARS-CoV-2. In this study, we analyzed aggregated foot traffic data in different place categories in NYC. Our study resulted in several findings that can inform improved neighborhood-level forecasting of disease outbreaks.

First, we identified differential mobility and indoor contact patterns in different place categories, likely influenced by the geographical distribution of POIs, the nature of human activities (e.g., dining versus shopping), and socioeconomic factors (e.g., working from home versus in an essential business) [52,53]. These differences explained heterogeneous disease burdens across NYC neighborhoods in our process-based model.

Second, aggregated foot traffic data for typical human activities are sufficient to support neighborhood-level infectious disease modeling without privacy concerns. A fundamental question in infectious disease modeling is whether the inclusion of more detailed processes results in more precise epidemic simulation, and to what extent system granularity is needed to inform real-world application of model outcomes. High-granularity epidemic models informed by human contact data are increasingly used in real-time forecasting. However, fine-grained contact data impose strong structural constraints on model dynamics, creating challenges in calibrating high-dimensional models to high-resolution real-world data [54]. The behavior-driven epidemic model, working at the neighborhood scale, provides a tradeoff between model realism, privacy protection, and computational efficiency.

Third, the inclusion of place category-specific FOIs in the behavior-driven epidemic model improved short-term forecasting at the neighborhood level. The more precise representation of infection risk in different place categories enhanced the forecast skills of the model. However, the proposed forecasting system, which assumes constant mobility and contact patterns in the forecast horizon, may not improve longer-term forecasts. This limitation underscores the necessity of predicting behavior changes in response to disease outbreaks and modeling the feedback loop between behaviors and epidemics. While theoretical work exists on this topic [55–62], empirical evidence of the impacts of policies and behaviors on epidemic spread needs to be quantified more precisely [63]. Currently, data-driven modeling applications and validations in real-world settings remain scarce.

While the behavior-driven epidemic model reproduced COVID-19 spread across NYC neighborhoods in 2020, process-based modeling beyond 2020 must additionally consider population immunity and virus evolution. Cumulative infections, vaccination, and immunity waning collectively shaped the immunological landscape upon which SARS-CoV-2 circulated after 2020 [64]. The continuous evolution of SARS-CoV-2 led to the emergence of new variants with varying transmissibility, immune escape capability, and disease severity [65,66]. Modeling these complex processes poses a significant challenge, especially at local scales that hold direct relevance to policymaking.

Several inference approaches were used in this study. For model fitting, we chose to use the Metropolis-Hasting MCMC to estimate parameters. Because the parameters *a* and *b* are power-law exponents in the FOI terms, the magnitudes of the other parameters are very sensitive to their values. This ultra-sensitivity creates estimation difficulty if we adjust *a* and *b* during MCMC updates. To circumvent this technical challenge, we instead used a model selection framework. Specifically, we fixed a combination of *a* and *b* in the model, performed MCMC to estimate other parameters, repeated this process for multiple (n=961) combinations of *a* and *b*, and determined which combination can explained the observation data. We then used a Bayesian approach to estimate the credible intervals of *a* and *b*. For forecasting, we chose the ensemble adjustment Kalman filter for two reasons. First, the EAKF is computationally more efficient and thus more suitable for real-time deployment to generate forecasts. Second, the EAKF can adjust model states sequentially using the most recent observations. This capability of online learning can capture temporal changes of parameters due to shifting policies and other factors.

In the behavior-driven model, we assumed all place categories share the same *a* and *b* parameters in the FOIs. In reality, the impact of crowdedness and dwell time on FOI might differ across place categories. Developing a model with separate *a* and *b* parameters in different settings is possible; however, estimating these additional parameters is challenging using passively collected foot-traffic data. Future studies may consider systematically collecting data in different place categories to quantify the differential dependency of FOIs on crowdedness and dwell time.

The MCMC model fitting underestimated the citywide incidence in summer 2020 (Fig. 3). The mismatch during this period with low incidence is potentially caused by the computation of log-likelihood in the MCMC algorithm. In the definition of observation error distribution, we set a minimum observation error variance 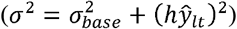, leading to a relatively larger uncertainty for low case numbers. Using this form, the log-likelihoods for high cases have more impact on the overall log-likelihood. As a result, the fitting will prioritize the performance during periods with high cases (i.e., the spring wave and winter wave). From a practical point of view, those periods are likely more important for disease control. The tradeoff between the high-case and low-case periods may have caused the systematic fitting bias in the summer of 2020. The underestimation during the summer was potentially forced by the model struggling to fit the downward trend of the spring wave while mobility was gradually increasing.

In retrospective forecasts, the behavior-driven model tended to overpredict COVID-19 cases in the summer of 2020. This was potentially driven by several factors – mobility, seasonality, and observation uncertainty. (1). The mobility in summer 2020 increased following the reopening of the city (Fig. 1A). This increased population mixing created the conditions for generating higher incidence in the model. (2). The seasonality forcing in the summer slightly reduced the transmission rate but did not offset the effect of increased mobility. (3). The ratio of uncertainty to incidence number during the summer was larger due to the minimum observation error variance imposed in the observation model. As a result, the update of model was less sensitive to low observed case numbers. The choice of observation error distribution may have contributed to the biased upward predictions in summer; however, the major driver was likely the increased mobility. This can be seen from the comparison with the baseline model B3, where the model has the same observation error distribution but static mobility matrices. Without mobility increase in the summer, model B3 did not generate overpredictions, suggesting that the EAKF was able to adjust the model in light of the consistently low observed incidence. While the behavior-driven model overestimated cases in summer 2020 in retrospective forecasts (Fig. S11), we argue that this forecasting bias does not lead to erroneous assessment of the general disease situation – the predicted case number is still very low compared to the spring wave. This comparison shows that the choice of mobility data (e.g., real-time versus historical average) in real-time forecasting should be made carefully.

The behavior-driven epidemic model, formulated in a metapopulation structure, may be deployed in real time to support outbreak response. As mobility and indoor contact patterns can concomitantly modulate the transmission of a range of respiratory pathogens, this model may be generalized to work for other respiratory pathogens sharing similar transmission routes with proper modifications. Several practical challenges still exist for real-time forecasting. First, epidemiological parameters for novel pathogens are difficult to estimate during early outbreaks. In a real-world setting, model parameters can be assigned using early estimates and subsequently updated when more data are accumulated. Sensitivity analyses on key parameters can also help to quantify uncertainty given imperfect selection of these parameters. Second, disease and mobility data released in real-time may be altered later to correct anomalies or underreporting. Such “backfill” makes real-time data less reliable when generating forecasts. Due to the lack of vintaged disease and mobility data that track these backfills, the behavior-driven model will likely perform less well in real-time applications than in retrospective forecasts in this study. In addition, the added value of incorporating mobility data into real-time forecasting depends on the latency and back-correction. Developing nowcasting approaches may help to alleviate the impact of backfill and improve the accuracy of real-time forecasts.

## Materials and Methods

### Data

Real-world weekly COVID-19 confirmed cases aggregated in 42 UHF neighborhoods were provided by the NYC DOHMH to support model fitting and retrospective forecasting. Case data in congregated settings were excluded to represent community transmission of SARS-CoV-2. We tracked mobility and contact patterns of NYC residents using aggregated foot traffic data covering 90,164 POIs. For each POI, the hourly number of visitors and home locations of visitors down to the census block groups level were recorded. To enhance privacy, SafeGraph excludes census block group information if fewer than five devices visited an establishment in a month from a given census block group. Other relevant metadata of POIs include the type of place (NAICS code), physical area (square meters), and daily average dwell time of visitors (minutes). We constructed the daily mobility matrices for each place category across the 42 neighborhoods by aggregating visitors to relevant POIs at the neighborhood level. For visitors without home information, we assigned their home neighborhoods using the distribution of visitors with home information in the same POI. Crowdedness for each POI was measured by dividing the daily visitor count by its physical area. Daily crowdedness and dwell time for each place category were averaged across all POIs within that category. Daily absolute humidity for NYC was derived from North American Land Data Assimilation System data [67].

### Modeling seasonality of SARS-CoV-2 transmission

COVID-19 cases, hospitalizations, and deaths can have both winter and summer peaks. This seasonality of COVID-19 infection is likely caused by the compound effects of the seasonality of the intrinsic transmissibility of the virus, population susceptibility, pathogen evolution, and length of immunity. In our model, we imposed a seasonal forcing of intrinsic transmissibility driven by absolute humidity. Below we discuss the impact of these factors on the seasonality of COVID-19 infections.

Many epidemiological studies have examined the impacts of weather and climate conditions on the seasonality of SARS-CoV-2 transmission. For instance, one study found that the transmissibility of SARS-CoV-2 in the US was modulated by meteorological factors including temperature, absolute humidity, and UV radiation, with absolute humidity playing a major role [38]. Another study also found that both temperature and absolute humidity have consistent negative effects on COVID-19 cases across both hemispheres [68]. While temperature in indoor settings is controlled and does not vary much over different seasons, indoor absolute humidity has a strong seasonality that is similar with outdoor environments [69]. These findings motivated use of absolute humidity to impose seasonal forcing on the transmissibility of SARS-CoV-2.

The emergence of summer peaks for COVID-19 infection may result from the dynamical interplay between virus transmission, population susceptibility, pathogen evolution, and duration of immunity. For instance, the summer peak in southern US states in 2020 was largely driven by high population susceptibility. Baker et al. [70] argued that susceptible supply limited the role of climate factors in the early SARS-CoV-2 pandemic. The summer wave in 2021 was likely caused by the emergence of the Delta variant. A study found that the Delta variant had a higher transmissibility compared to the ancestral SARS-CoV-2 strain and was capable of immune escape [71]. Later, the Omicron variant and its subvariants continued to emerge, with substantial immunity escape capability [72]. Many studies found that the duration of immunity to Omicron from prior infection wanes quickly. A review reported that the effectiveness against reinfection of protection induced by prior Omicron infection waned to 24.7% (95% CI 16.4–35.5) at 12 months [73]. Such fast waning of immunity can accumulate enough susceptible population within a year and produce summer peaks of COVID-19 infection in 2023 and beyond.

To examine whether the inclusion of seasonal forcing improves model performance, we generated retrospective forecasts using a baseline model without the seasonality term (Model B2). We found that the performance of this model was inferior to the model with humidity-forcing (Fig. 4).

### Model configuration and observation model

The behavior-driven epidemic model was initiated on February 26, 2020, prior to the reporting of the first COVID-19 case in NYC on March 1, 2020. Initial infections on the first day of simulation were informed by estimated infections that accounted for the severe underreporting during the early phase of the pandemic [74]. Specifically, we randomly drew initial infection numbers in NYC from a uniform distribution *U*[500,20000] and distributed them to the 42 neighborhoods as infectious population (*I*) in the model according to the distribution of cumulative cases reported within three weeks after March 1, 2020. The exposed population (*E*) in each neighborhood was set as *E*= γ*I* ([γ ∈*U*[2,6]), representing rapid increase of infections before the detection of local cases. The remainder of the population was set as susceptible (*S*). Epidemiological parameters (*z* and *d*) were fixed to *z* = 3.59 days and *d* = 3.56 days. The incubation and infectious periods were estimated using early infection data from the US [75]. The seasonality parameters were set as *w*_*max*_ = 2.6 and *w*_*min*_ = 1.4, informed by the rough estimate of the basic reproductive number of SARS-CoV-2 in winter and summer. The seasonality of virus transmissibility was modulated by the daily absolute humidity data in NYC. Sensitivity analysis was performed for other values of *w*_*max*_ and *w*_*min*_ (*w*_*max*_ ∈ {2.5,2.6,2.7,2.8,2.99} and *w*_*min*_ ∈ {1.1,1.2,1.3,1.4,1.59}) and the performance of fitting was robust.

Daily confirmed cases were computed using an observation model. To account for varying surveillance efforts across communities and over time, we used the weekly ascertainment rates estimated by Yang et al. [42] for each neighborhood validated by case, hospitalization, death, serological, and wastewater data (Fig. S3). We further introduced a gamma distribution Γ (1.85,7.57) to model the delay from infection to case conformation, similar to our prior works [9,46,47]. Among the newly exposed population *E*_*new*_ (*t*) on a given day *t, α* fraction will be reported. These new infections will be confirmed on day *t*+ *t*_*d*_, where *t*_*d*_ is drawn from the gamma distribution.

### Markov Chain Monte Carlo and model fitting

We fit the model to weekly confirmed cases in 42 neighborhoods using Metropolis-Hastings (MH) MCMC. For each combination of parameters *a* and *b*, we performed MCMC independently. Within each MCMC fitting, *v*_*p*_ for five place categories, *β*_*h*_ for baseline transmission, and a parameter controlling for the magnitude of observation error (see details below) were estimated and other epidemiological and seasonality parameters were fixed as described in model configuration. The goal of MCMC is to estimate the distributions of model parameters ***x*** = (*v*_1_, *v*_2_,*v*_3_, *v*_4_, *v*_5_, *β* _*h*_, *h*)^*T*^ given observed case data ***O***: *P* (***x***|***O***) ∝ P(*x*)*P* (*0*|*x*), where *P*(*x*) is the prior distribution and *P*(***O***|***x***) is the likelihood of observing ***O*** given model parameters ***x***. The Metropolis-Hasting algorithm proceeds as follows:

1. Initialization. Select initial states of unknown parameters 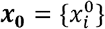 from prior distributions, where 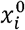 is the initial value of the *i*^th^ parameter.
2. Iteration. For each parameter 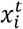 at iteration *t*, sequentially perform the following procedures while fixing other parameters at their current states.
  a. Generate a random candidate state 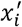 according to a proposal distribution 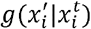.
  b. Calculate the acceptance probability 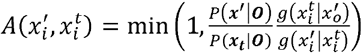 where *x*_*t*_ is the current model parameter vector and *x*′ is the perturbed vector with 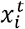 replaced by 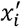.
  c. Accept or reject the candidate state 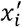. Draw a uniform random number *u*∈ *U*[0,1]. If 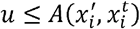, update 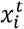 to 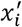; otherwise, keep 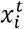 unchanged.
  d. After looping through all parameters, move to the next iteration *t* = *t* + 1.

In MCMC, the prior distributions of parameters were set broadly as *v*_*p*_ ∈ *U*[0,1000], *β*_*h*_ ∈ *U*[0,0.50], and *h*∈ *U*[0.1,10]. The ranges of these parameters were explored and selected to ensure that the model can produce outbreaks with a similar scale as in the real-world data. The proposal distributions were symmetric. For each parameter, we perturbed the current parameter value by adding Gaussian random noise with a standard deviation 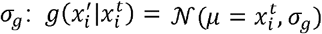. Because the parameters have different scales and the observations have different sensitivities to these parameters, we selected *σ*_*g*_ heuristically for each parameter: *σ*_*g*_ is 5, 5, 10, 15, 5 for each *v*_*p*_, 0.02 for *β*_*h*_, and 0.1 for. We next compute the acceptance probability 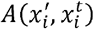. As the proposal distributions are symmetric, the term 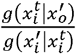 equals 1. We further have 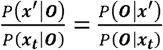, which is the ratio of the two likelihoods. Denote *P* (***O***|***x***) as exp(𝓁 (***x***)), where 𝓁 (***x***) = log *P*(***O***|***x***) is the log-likelihood given the parameter vector *x*. The acceptance rate can be written as the 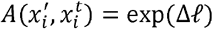, where Δ𝓁 = 𝓁 (***x**′*) - 𝓁 (***x***_***t***_) is the difference of log posterior after parameter update. Intuitively, if the parameter update does not decrease the log posterior (i.e., Δ𝓁 ≥ 0), the candidate parameter is accepted; otherwise (i.e., Δ𝓁 < 0), the candidate parameter is only accepted with a probability that decreases exponentially with the change of log posterior.

To compute the log-likelihood, we assumed Gaussian observation errors in the model. Note, discrete, positive-valued distributions such as the negative binomial and Poisson distributions are suitable to model infection data, particularly for low incidence. Here, the choice of the Gaussian observation errors is to keep the observation noise form consistent with the EAKF in retrospective forecasting, where the likelihood of observation is assumed to be Gaussian. For the EAKF, both the prior and likelihood are assumed to be normally distributed, so that the posterior distribution is also normal. This choice is partly due to mathematical convenience and tractability. In practice, it also has worked well in previous applications to infectious disease forecasting [40]. For each set of parameters, we ran the epidemic model throughout the fitting period to generate weekly reported cases in all neighborhoods. The log-likelihood of observing real-world weekly case data was approximated as 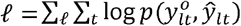, where the summation ran over all neighborhoods and weeks. The term 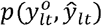 is the probability of the observation 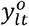 for a Gaussian distribution 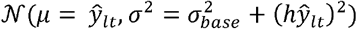. Here, 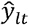 is the simulated weekly cases in location *l* at week *t, σ*^2^ is the observation error variance, 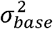 is the minimum observation error variance (set as 20^2^), and *h* is a parameter controlling the relationship between *σ*^2^ and 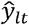. This heuristic form ensures that the variance *σ*^2^ has a minimum baseline value for zero cases and that the standard deviation *σ* increases (almost) linearly with case number for large observation values. Here the baseline variance was set as 20^2^ heuristically but can be adjusted based on model performance. To estimate the Gaussian error, we included the parameter *h* in the MCMC fitting.

We fit the model to weekly neighborhood-level COVID-19 cases from March 1, 2020 to December 13, 2020. We ran 1 million MCMC steps and estimated the posterior parameter distributions using 10 thousand samples taken every 95 steps from the last 950 thousand steps. We used trace plots, Gelman-Rubin 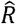 statistics, and effective sample size to examine the convergence of MCMC chains (see Fig. S5). The Gelman-Rubin 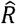 statistics for the six parameters (*v*_1_*v*_2_ *v*_3_*v*_4_*v*_5_, *β*_*h*_,*h*)^*T*^ are 1.10, 1.01, 1.08, 1.01, 1.00, 1.06, and 1.02. The effective sample sizes (percentage of posterior samples) are 33.5%, 34.5%, 33.4%, 41.7%, 35.7%, 33.5%, and 64.7%. We performed four independent MCMC chains starting from randomly selected initial infections in NYC from *U*[500,20000]. The convergence of several parameters was not ideal based on the Gelman-Rubin 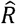 statistics. This is possibly caused by the correlation between mobility patterns across place categories, particularly the synchronous drop during the lockdown. While merging place categories can potentially improve the convergence, we chose to keep the current classification for explicit representation of different activities. Ideally, if we have access to mobility data with stronger variation across place categories and associated disease data, it is possible to more accurately estimate model parameters with better convergence. Future works can be done to collect more informative data to estimate parameters. We note that more sophisticated and efficient MCMC methods such as the No-U-Turn Sampler [76] can explore complicated posterior distributions and provide more informative evidence of non-convergence [77]. These methods can be tested in future studies. In addition, in model fitting, we found that the magnitude of the posterior parameters *v*_*p*_ was sensitive to the choice of power exponents *a* and *b*. We speculate that the sensitivity of posterior parameters *v*_*p*_ was caused collectively by the functional form of FOIs and the parametric flexibility to find the most useful proxies for transmission in mobility data. This suggests the need to perform controlled experiments to better estimate the parameters *a* and *b*.

### Retrospective forecasts and evaluation metrics

We used the model-data assimilation (M/D/A) framework to generate retrospective forecasts. This framework has been widely used in numerical weather forecasting and infectious disease forecasting [40,78–80]. Specifically, we coupled the behavior-driven epidemic model with an efficient data assimilation algorithm, the ensemble adjustment Kalman filter (EAKF) [45]. The forecasting framework was assumed to be the following: we first used data from the spring wave (until June 7, 2020) to estimate key parameters such as *a* and *b*, and then used the model state on June 7, 2020 to initiate the forecasting from June 8, 2020. In the EAKF, an ensemble of model states was used to represent distributions of model variables and parameters. In our implementation, 500 ensemble members were used. For each ensemble member, we randomly drew the parameters *a* and *b* from their posterior distributions to capture their uncertainty and set *h*= 0.40 (the posterior mean of *h* in the training period up to June 7, 2020), fixing these parameters during the forecast. Each week, parameters *v*_*p*_ for five place categories and *β* _*h*_ for baseline transmission, were updated so that the model could better fit the most recent observation. The observed model states (weekly confirmed cases) were updated using the Bayes’ rule and unobserved model states were updated using their cross-ensemble covariability with observed model states. Forecasts were generated using posterior parameters and variables. We assumed a constant mobility and contact patterns during the short-term forecast horizon. To avoid negative values in predictive distributions, we clipped the Gaussian observation distribution and set negative values to be 0. For parameter values that went outside the prior range in the EAKF update, we also applied a clipping to set them at the boundaries of the prior range. More implementation details and the pseudo-code of the EAKF are provided in Supplementary Information.

The forecast system generates probabilistic predictions. We evaluated the forecast skills using four metrics: (1) mean absolute percentage error (MAPE), (2) log score, (3) weighted interval score (WIS) [48], and (4) coverage of 95% predictive intervals. For a weekly forecast in a neighborhood, MAPE is defined as 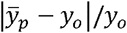, where 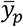 is the mean prediction of weekly case and *y*_*o*_ is the observed value. To compute the log score, we first applied a kernel density estimation to fit the predictive distribution (represented by the discrete ensemble members) and took the logarithmic value (base *e*) of the probabilistic density at the observation. WIS is a proper scoring rule for quantile forecasts, which was recently used in evaluating probabilistic predictions of infectious diseases [50]. It converges to the CRPS (continuous ranked probability score) for an increasing number of intervals. We followed the configurations used by the CDC COVID-19 forecast hub and the FluSight challenge, using 23 quantiles (0.01, 0.025, 0.05, 0.1, …, 0.95, 0.975, 0.99) estimated from the 500 ensemble members. The WIS scores were computed using the R function provided at https://github.com/cmu-delphi/covidcast. The coverage for 95% predictive intervals was computed as the fraction of real-world observations falling within the 95% predictive intervals over the entire forecasting period. The coverage of a perfectly calibrated forecast should be 0.95 for 95% predictive intervals. Given the wide range of the forecast targets, future work may consider using log-transformed case incidence as a more appropriate forecast quantity to score. Scoring on the log-transformed scale may better reward accurate growth rate inference and prediction [44].

### The baseline forecast models

We generated retrospective forecasts using three baseline models: (B1), a metapopulation model without place category-specific mobility but with seasonal forcing; (B2), the behavior-driven model without seasonal forcing; (B3), the behavior driven model with seasonal forcing but static mobility matrices, crowdedness, and dwell time, averaged over the week prior to June 8, 2020. Specifically, the model dynamics of B1 is described by the following equations.

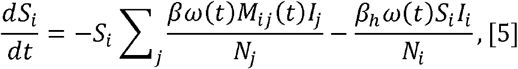

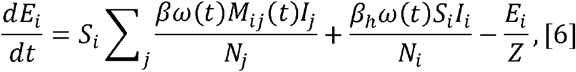

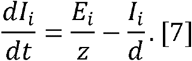

Here *N*_*i*_, *s*_*i*_, *t*_*i*_, and *I* _*i*_ are the total, susceptible, exposed, and infectious population in neighborhood *i*; *z* and *d* are the latency and infectious duration; *β* is the transmission rate in POIs captured by the foot traffic data; *β* _*h*_ is the baseline transmission rate in other uncaptured places; *ω* (*t*)is seasonality term forced by absolute humidity; and *M*_*ij*_ (*t*)represents the fraction of population living in neighborhood *i* visiting neighborhood *j* on day *t*, computed using the same foot traffic data. Note, the baseline model incorporated time-varying cross-neighborhood mobility but did not include the place category-specific FOIs informed by crowdedness and dwell time. The transmission dynamics of B2 is described by

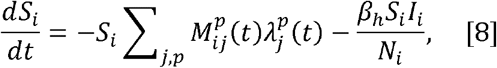

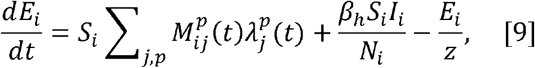

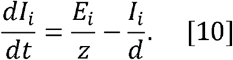

The model dynamics of B3 is described by

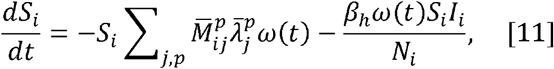

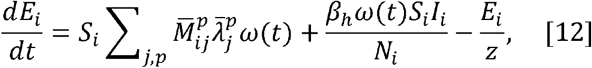

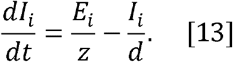

Here 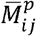 is the static mobility matrix and 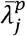 is the static FOIs using crowdedness and dwell time averaged the week prior to June 8, 2020.

## Data Availability

The aggregated neighborhood-level foot traffic data, climate data, and codes for the behavior-driven epidemic model, model fitting, and retrospective forecasting are shared publicly at GitHub (https://github.com/LydiaTai/Behavior-driven-forecast). The neighborhood-level COVID-19 case data in NYC are available from NYC DOHMH, but restrictions apply to the availability of these data, which were used under license for the current study, and so are not publicly available. Requests for data access should be addressed to NYC DOHMH at. COVID-19 data in NYC can be found at https://www.nyc.gov/site/doh/covid/covid-19-data.page. Synthetic outbreak data at the neighborhood level are shared as an example to run the analysis at the GitHub repository.

## Supporting information

**S1 Text. Supplementary materials**. Extended methods and analyses. (PDF)

