## Supplementary Materials for "Behavior-driven forecasts of neighborhood-level COVID-19 spread in New York City"

**S1 Text**

Defining place categories

To examine whether the same type of POIs shared similar visitation and contact patterns, we performed a clustering analysis of POIs based on three metrics: (1) crowdedness, (2) dwell time, and (3) variance of visitor numbers within a week. We first classified all POIs into subgroups using the first four digits of the NAICS code. For each subgroup, we calculated the mean values of all three metrics using data from January 2020 to February 2020 (prior to the pandemic). We normalized each metric by dividing the maximum value across all subgroups so that the three metrics are on the same scale. A Gaussian mixture clustering was performed. We identified three clusters (Fig. S1). Interestingly, subgroups of POIs within the same broader categories were generally clustered within the same cluster. Based on this preliminary analysis, we classified all POIs into five place categories using the first four digits of NAICS codes. The classification rule is provided in Table S1.

Estimating the dependency of FOIs on crowdedness and dwell time

We used a Bayesian approach to approximate the uncertainty of parameters $a$ and $b$. We use $\theta$ to denote all model parameters other than $a$ and $b$ and $y$ to denote observation data. Assume the priors for $(a, b)$ are independent from the priors for $\theta$. The marginal posterior for the parameters $a$ and $b$ is:

$$\begin{aligned} p(a,b|y) \propto p(a,b)\cdot p(y|a,b) &= p(a,b)\cdot \int p(y,\theta|a,b)d\theta \\ &= p(a,b)\cdot \int p(y|\theta,a,b)p(\theta|a,b)d\theta \\ &= p(a,b)\cdot \int p(y|\theta,a,b)\frac{p(\theta)}{p(\theta|y,a,b)}p(\theta|y,a,b)d\theta. \end{aligned}$$

The likelihood $p(y|a,b)$ can be approximated by

$$\frac{1}{n}\sum_i p(y|\theta_i,a,b)\frac{p(\theta_i)}{p(\theta_i|y,a,b)},$$

where $\theta_i, i = 1,\cdots,n$ are drawn from the posterior for $\theta$ given the fixed values of $a$ and $b$. Since $\theta$ includes seven parameters, directly estimating this high-dimensional distribution is challenging. We therefore used a crude approximation. For each combination of $(a, b)$, we used the samples of $\theta_i$ from the MCMC chain (1000 samples taken every 5 steps from the last 5000 steps) to approximate the posterior of $p(\theta_i|y,a,b)$. Using the corresponding likelihood $p(y|\theta_i,a,b)$ for each sample $\theta_i$ in the chain, we computed the likelihood $p(y|a,b)$ for discrete combinations of $(a, b)$ using the above equation. Note in this approximation, we assumed the ratio of the prior and the posterior distributions is equal to one, which could introduce approximation errors. Now, for each combination of $(a, b)$ we tested in the grid search, we obtained the estimation of likelihood $p(y|a,b)$. We fit the log-likelihood surface using a cubic spline interpolation and normalized the likelihood obtained from the fitting. Assuming a uniform prior for $a$ and $b$ in [0,3], the posterior distributions of these parameters are proportional to their likelihoods. We then estimated the marginal posterior for each parameter by summing across the axis of the other parameter.

Retrospective forecasts

We used the EAKF to generate retrospective forecasts. The EAKF assumes a Gaussian distribution of both the prior and likelihood and adjusts the prior distribution to a posterior using Bayes' rule deterministically. To represent the state-space distribution, the EAKF maintains an ensemble of system state vectors acting as samples from the distribution. In particular, the EAKF assumes that both the prior distribution and likelihood are Gaussian, and thus can be fully characterized by their first two moments (mean and variance). The update scheme for ensemble members is computed using Bayes' rule (posterior $\propto$ prior $\times$ likelihood) via the convolution of the two Gaussian distributions. For observed state variables, the posterior of the $i$th ensemble member is updated through

$$y_{t,post}^{i} = \frac{\sigma_{t,obs}^{2}}{\sigma_{t,obs}^{2} + \sigma_{t,prior}^{2}} \bar{y}_{t,prior} + \frac{\sigma_{t,prior}^{2}}{\sigma_{t,obs}^{2} + \sigma_{t,prior}^{2}} y_{t}^{o} + \sqrt{\frac{\sigma_{t,obs}^{2}}{\sigma_{t,obs}^{2} + \sigma_{t,prior}^{2}}} \left(y_{t,prior}^{i} - \bar{y}_{t,prior}\right). \quad [S1]$$

Here $y_{t,post}^{i}$ and $y_{t,prior}^{i}$ are the posterior and prior of the observed variable for the $i$th ensemble member at time $t$; $\bar{y}_{t,prior}$ is the mean of the prior observed variable; $\sigma_{t,obs}^{2}$ and $\sigma_{t,prior}^{2}$ are the variances of the observation and the prior observed variable; and $y_{t}^{o}$ is the observation at time $t$. Unobserved variables and parameters are updated through their covariability with the observed variable, which can be computed directly from the ensemble. In particular, the $i$th ensemble member of unobserved variable or parameter $x^{i}$ is updated by

$$x_{t,post}^{i} = x_{t,prior}^{i} + \frac{\sigma\left(\left\{x_{t,prior}\right\}_{n}, \left\{y_{t,prior}\right\}_{n}\right)}{\sigma_{t,prior}^{2}} \left(y_{t,post}^{i} - y_{t,prior}^{i}\right). \quad [S2]$$

Here $x_{t,post}^{i}$ and $x_{t,prior}^{i}$ are the posterior and prior of the unobserved variable or parameter for the $i$th ensemble member at time $t$; and $\sigma\left(\left\{x_{t,prior}\right\}_{n}, \left\{y_{t,prior}\right\}_{n}\right)$ is the covariance between the prior of the unobserved variable or parameter $\left\{x_{t,prior}\right\}_{n}$ and the prior of the observed variable $\left\{y_{t,prior}\right\}_{n}$ at time $t$. In the EAKF, variables and parameters are updated deterministically such that the higher moments of the prior distribution are preserved in the posterior.

In the EAKF, we assumed a heuristic form of observation error variance (OEV): $\sigma_{lt}^{2} = \sigma_{l0}^{2} + (hy_{lt}^{o})^{2}$, where $\sigma_{l0}^{2}$ is the baseline OEV for neighborhood $l$ and $y_{lt}^{o}$ is the observed case in neighborhood $l$ in week $t$. To account for heterogeneous disease burdens across neighborhoods, the baseline OEV $\sigma_{l0}^{2}$ was defined as 400 (i.e., standard deviation 20). The parameter $h$ was fixed as 0.40 (the posterior mean of $h$ in the training period up to June 7, 2020). We chose a relatively large OEV to avoid ensemble degeneracy. Similar forms of OEV have been successfully used for inference and forecasting for a range of infectious diseases. During the weekly EAKF update, we looped through observations in all neighborhoods. The observation in neighborhood $l$ was used to update model variables within the same neighborhood and global parameters ($v_{p}$ for five place categories and $\beta_{h}$ for transmission not accounted for by mobility data). In implementation, we used 500 ensemble members.

A pseudo-code for the EAKF is provided below.

| Algorithm 1. EAKF |
| --- |
| **Input:** The transmission model $\mathcal{M}$, observations $\{y_{lt}^{o}\}$ in $T$ weeks and $L$ locations, the observational error variance (OEV) $\{\sigma_{lt}^{2}\}$, the initial ensemble of system state $\{z_{i}^{0}\}$ with 500 ensemble members. |
| **for** $t = 1$ to $T$ **do** |
| Run model $\mathcal{M}$ forward for one time step using system state $\{z_{i}^{t-1}\}$, obtain the ensemble of prior observed variables and system states at time $t$: $\left[\{y_{lt,prior}\},\{z_{i,prior}^{t}\}\right] = \mathcal{M}(\{z_{i}^{t-1}\})$ |

**for** $l = 1$ to $L$ **do**

Update observed variable in location $l$ at time $t$ using Eq. S1:

$$\{y_{lt,post}\} = EAKF(\{y_{lt,prior}\}, y_{lt}^{o}, \sigma_{lt}^{2}).$$

Update unobserved variables in location $l$ at time $t$ and parameters using Eq. S2:

$$\{z_{i,post}^{t}\} = EAKF(\{z_{i,prior}^{t}\}, \{y_{lt,prior}\}, \{y_{lt,post}\}).$$

**end for**

Update the ensemble of model state at time $t$ using the posterior: $\{z_i^t\} \leftarrow \{z_{i,post}^t\}$, which will be used to initiate the model at the next time step.

Run the transmission model $\mathcal{M}$ forward using current posterior state to generate forecasts.

**end for**

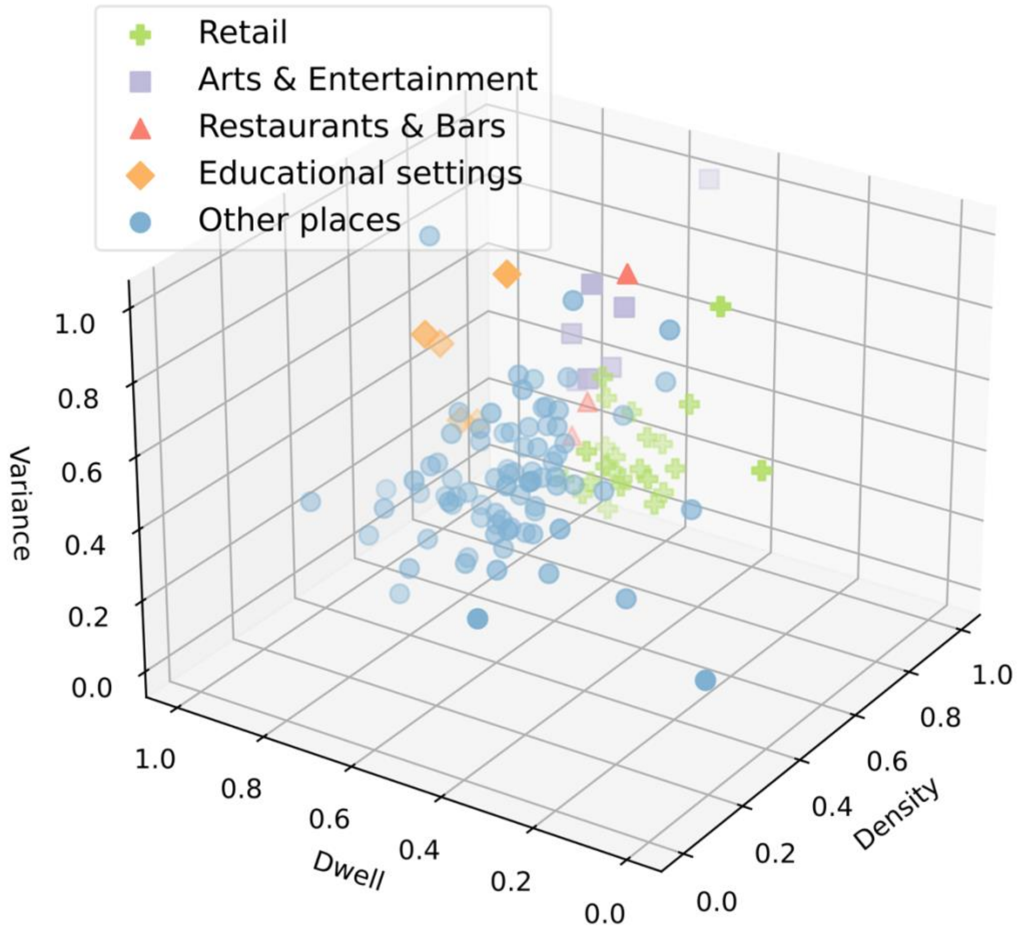


**Fig. S1. Clustering of POIs using crowdedness, dwell time, and variance of visitor numbers within a week.** Each data point represents one subgroup of POIs with the same first four digits of the NAICS code. Different place categories are represented using different colors and shapes.

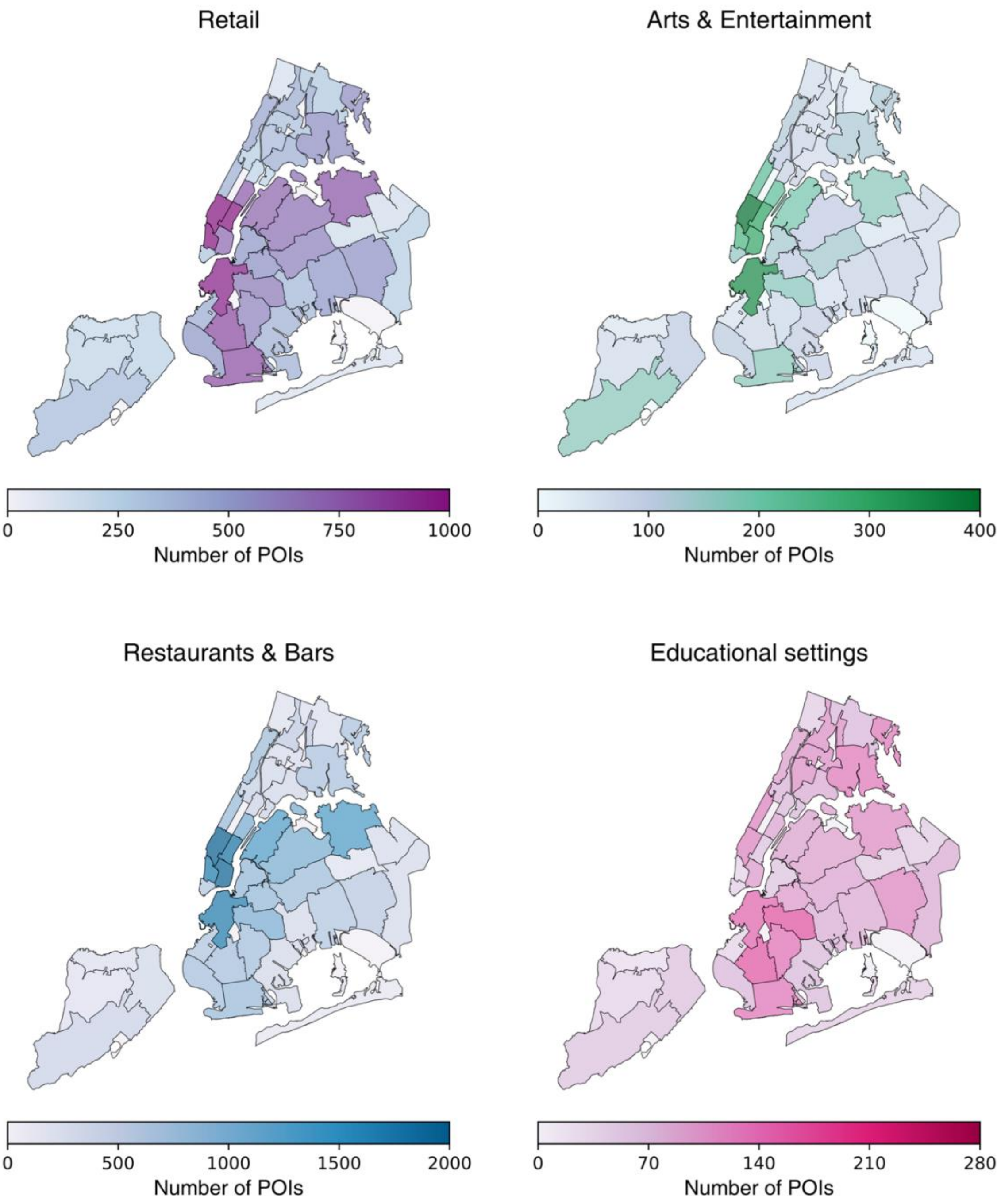


**Fig. S2. Geographical distribution of POIs.** We visualize the number of POIs in each neighborhood for retail, arts & entertainment, restaurants & bars, and educational settings. The maps were created using Python using the shapefile publicly available at https://github.com/nychealth/coronavirus-data/tree/master/Geography-resources. This is a public repository by NYC Department of Health and Mental Hygiene. The term of use can be found here: https://github.com/nychealth/coronavirus-data.

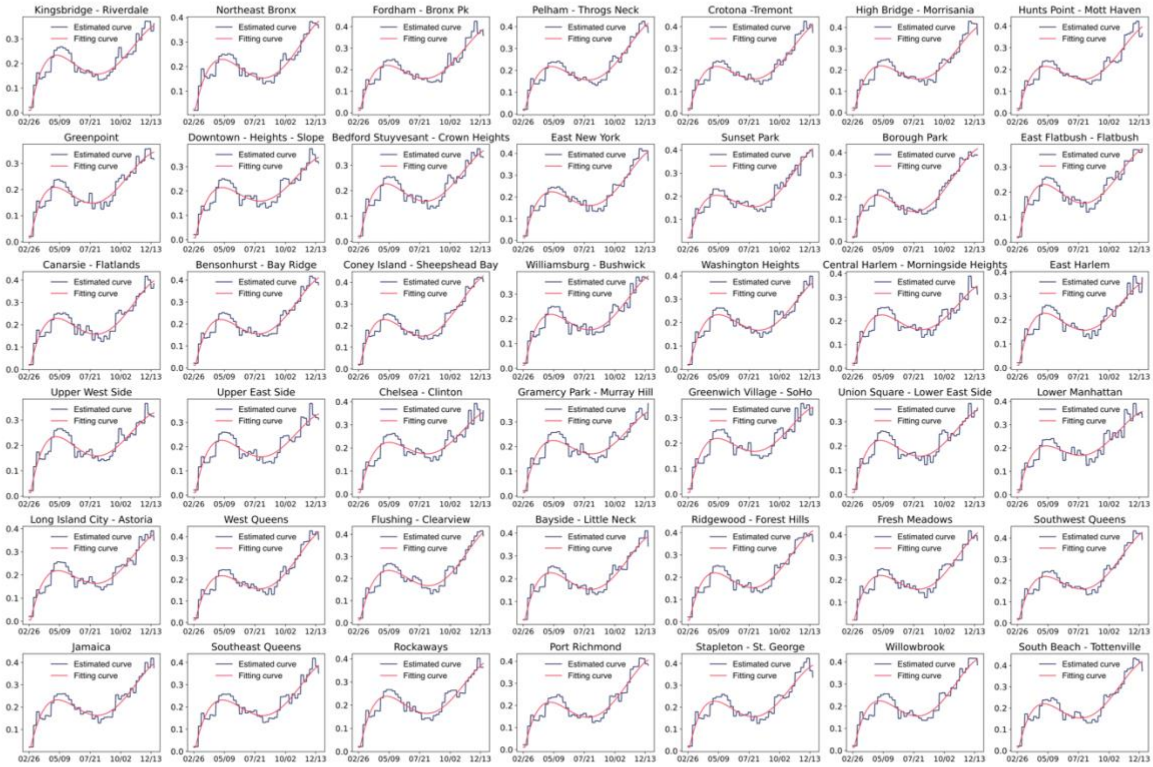


**Fig. S3. Weekly ascertainment rates in each neighborhood.** The black lines are the estimated weekly ascertainment rate in Yang et al 2022. We fit a smooth curve using a fourth-order polynomial function (red lines) for each neighborhood and used the smoothed curve to set ascertainment rates in our model.

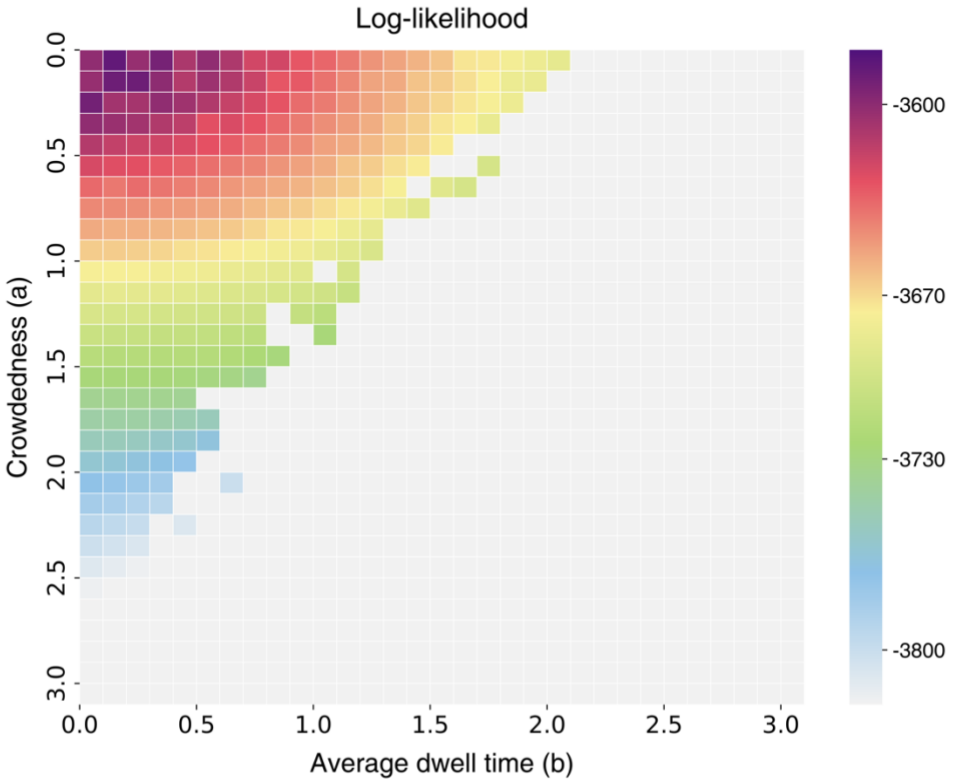


**Fig. S4. The log-likelihood of model fitting for combinations of $\boldsymbol{a}$ and $\boldsymbol{b}$.** The log-likelihood landscape for different combinations of parameters $\boldsymbol{a}$ (the power exponent for crowdedness) and $\boldsymbol{b}$ (the power exponent for average dwell time). We performed a grid search of $\boldsymbol{a}$ and $\boldsymbol{b}$. For each combination, we ran MCMC fitting and show the log-likelihood for the model fitting.

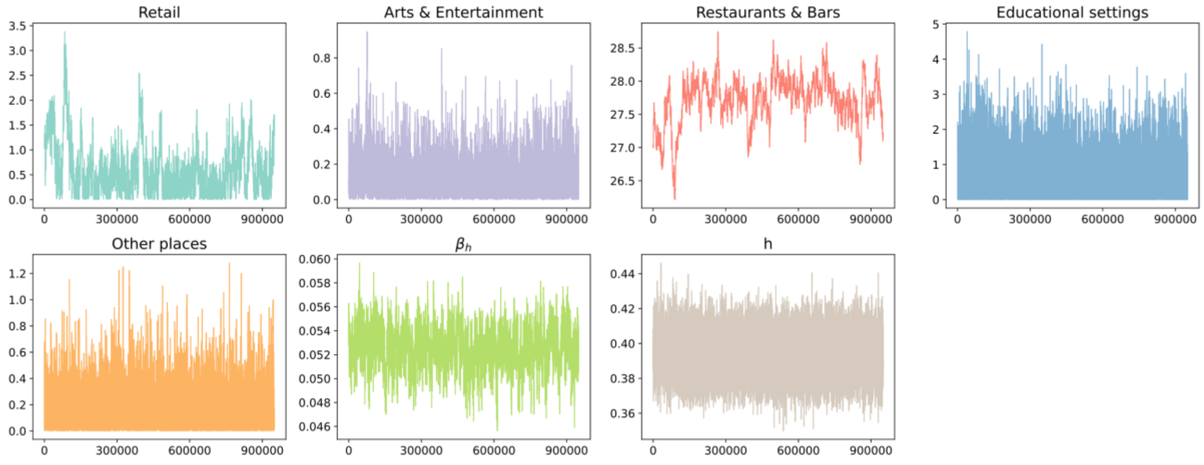


**Fig. S5. Trace plots for parameters in the MCMC.** We fit the behavior-driven model to the neighborhood-level data from March 1, 2020 to December 13, 2020, fixing the parameters $a = 0.30$ and $b = 0.13$. We ran the MH-MCMC algorithm for 1 million steps and plot samples in the latter 950,000 steps (excluding the first 50,000 steps as burn-in).

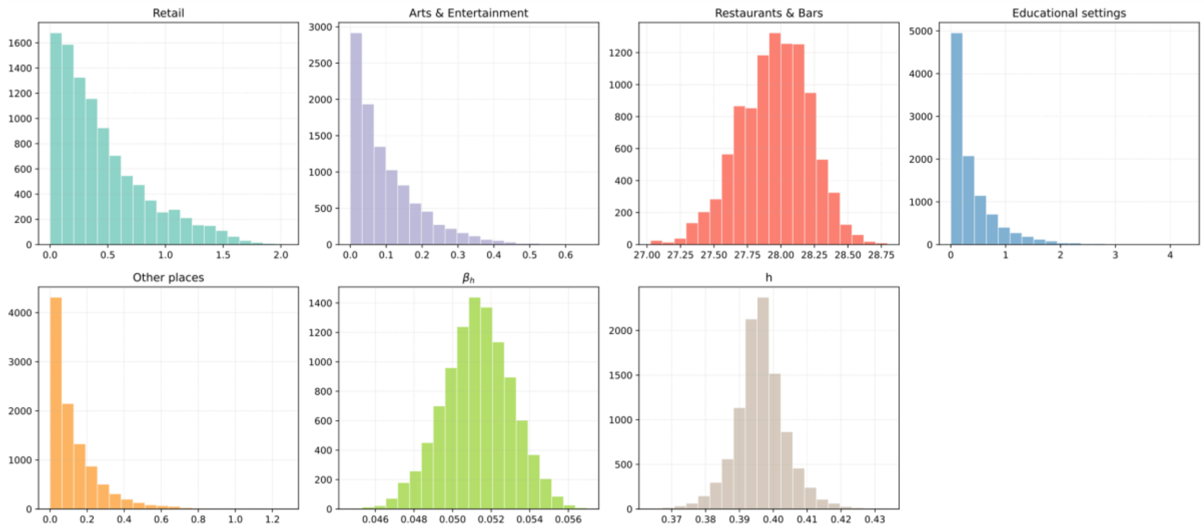


**Fig. S6. Posterior distributions of parameters.** We fit the behavior-driven model to the neighborhood-level data from March 1, 2020 to December 13, 2020, fixing the parameters $\boldsymbol{a} = \mathbf{0.30}$ and $\boldsymbol{b} = \mathbf{0.13}$. We ran the MH-MCMC algorithm for 1 million steps and discarded the first 50,000 steps as burn-in. The posterior distributions of parameters were estimated using 10,000 samples selected every 95 steps in the latter 950,000 iterations.

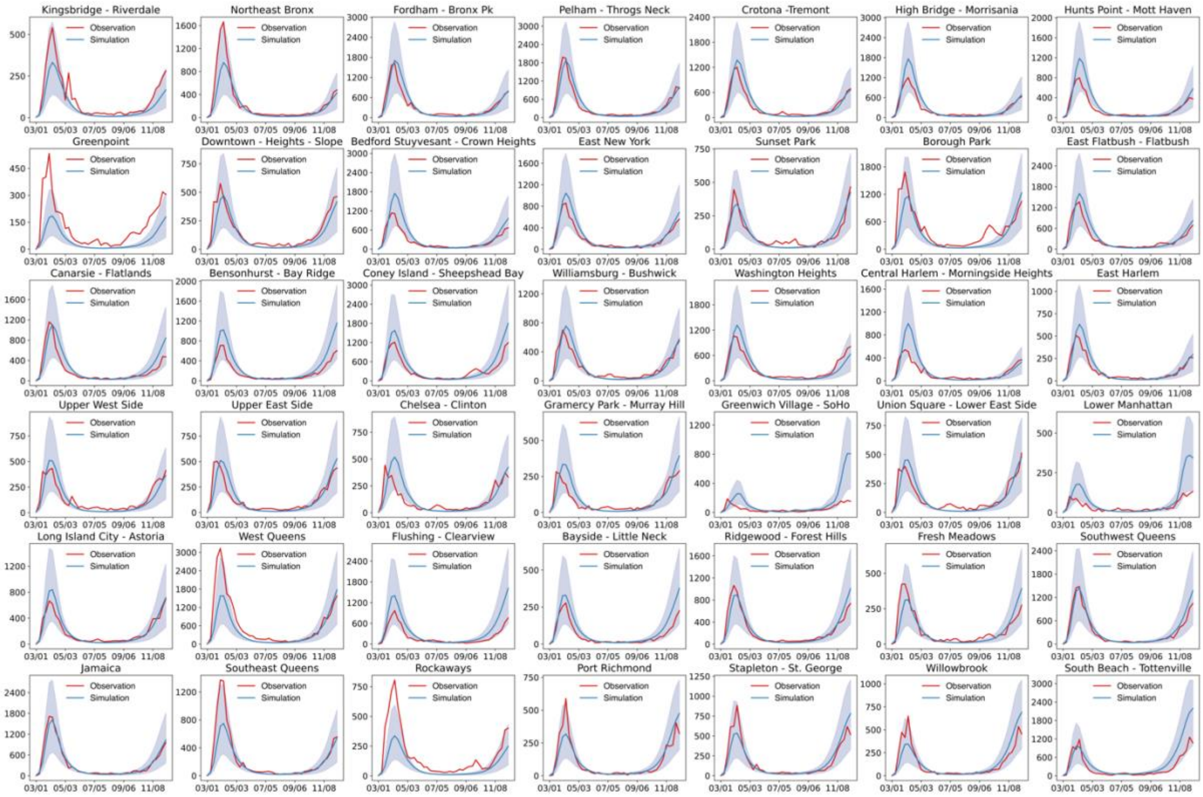


**Fig. S7. Model fitting in 42 NYC neighborhoods.** Model simulations using estimated posterior parameters (blue) are compared with the reported weekly cases in each neighborhood. Red lines are the observed weekly cases in NYC neighborhoods. The blue shaded areas show 95% CIs, obtained from 500 independent simulations, without adding the Gaussian observation error. The solid blue lines show the medians of 500 simulations.

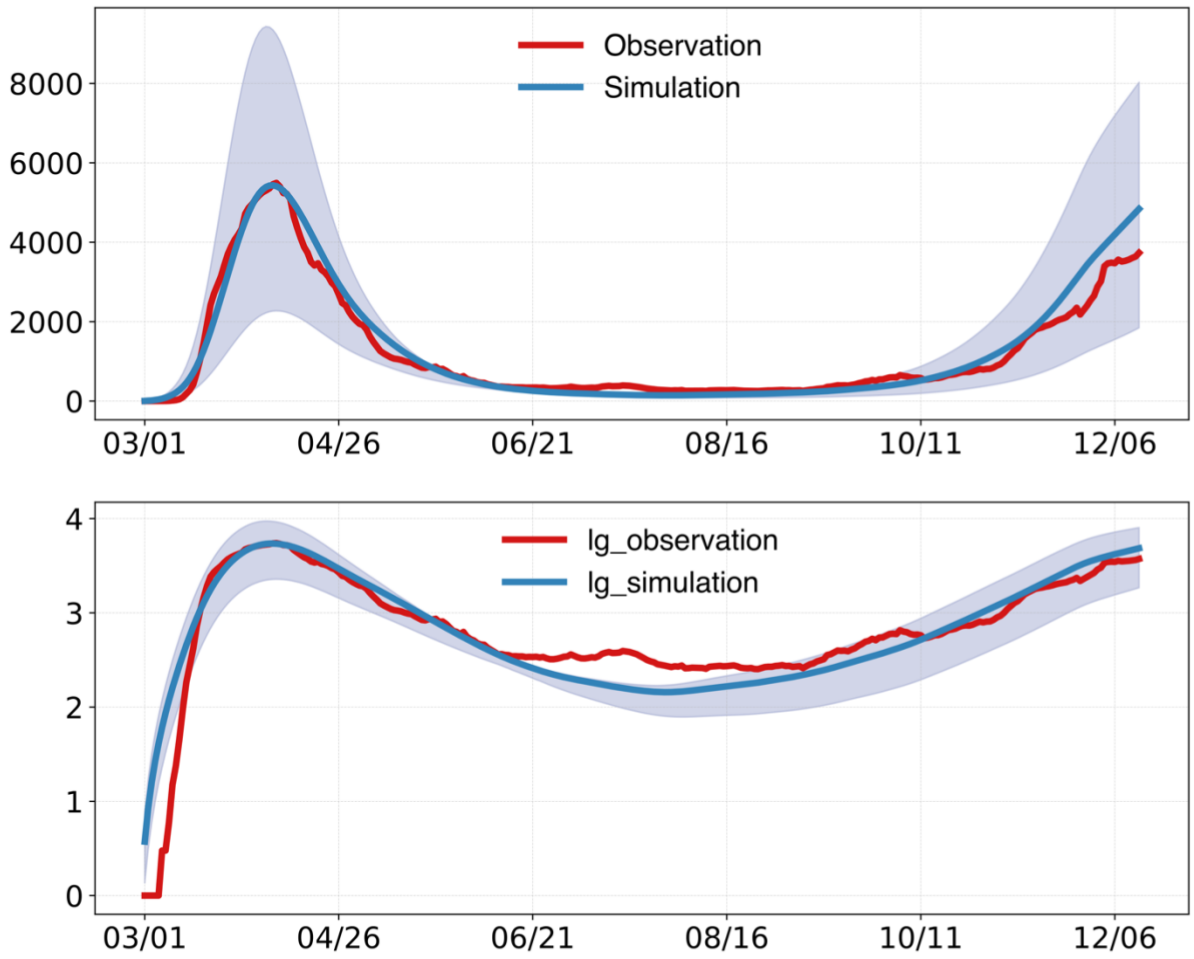


**Fig. S8. Model fitting for NYC using the behavior-driven model.** Simulations using model parameters estimated for the period from March 1, 2020 to December 13, 2020. Simulated cases were aggregated to the city level and are compared with the daily confirmed cases in NYC. The upper plot shows the fitting in a linear scale and the lower plot shows the fitting in a log scale. The shaded blue area represents the 95% credible interval, obtained from 500 independent simulations, without adding the Gaussian observation error (i.e., it only reflects the uncertainty in the latent deterministic trajectories). The solid lines represent the median of those 500 trajectories.

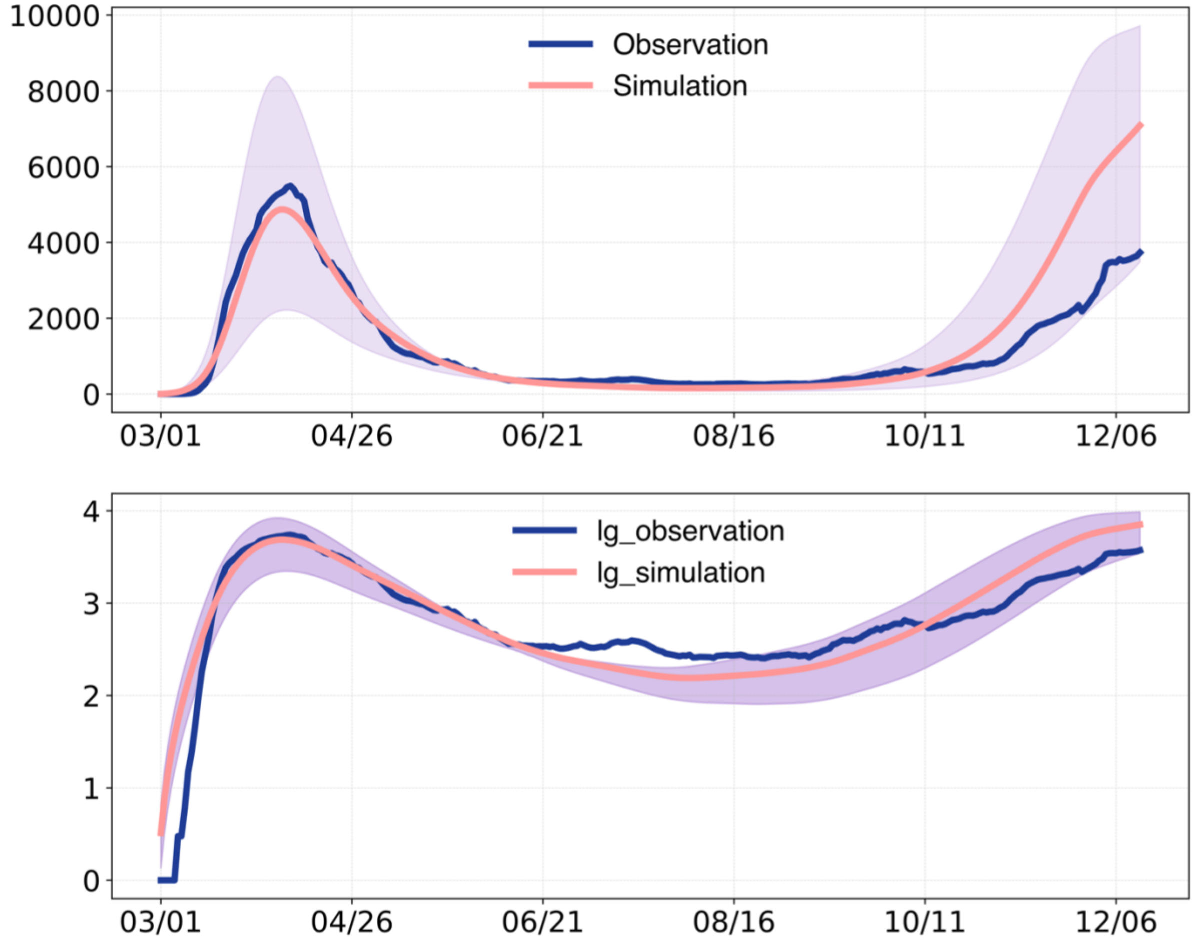


**Fig. S9. Model fitting for NYC using the behavior-driven model without crowdedness and dwell time.** We ran MH-MCMC algorithm to fit the neighborhood-level COVID-19 data using the behavior-driven model without crowdedness and dwell time (fixing $\boldsymbol{a} = \mathbf{0}$ and $\boldsymbol{b} = \mathbf{0}$). Simulations using model parameters estimated for the period from March 1, 2020 to December 13, 2020. Simulated cases were aggregated to the city level and are compared with the daily confirmed cases in NYC. The upper plot shows the fitting in a linear scale and the lower plot shows the fitting in a log scale. The shaded blue area represents the 95% credible interval, obtained from 500 independent simulations, without adding the Gaussian observation error. The solid lines represent the median of those 500 trajectories.

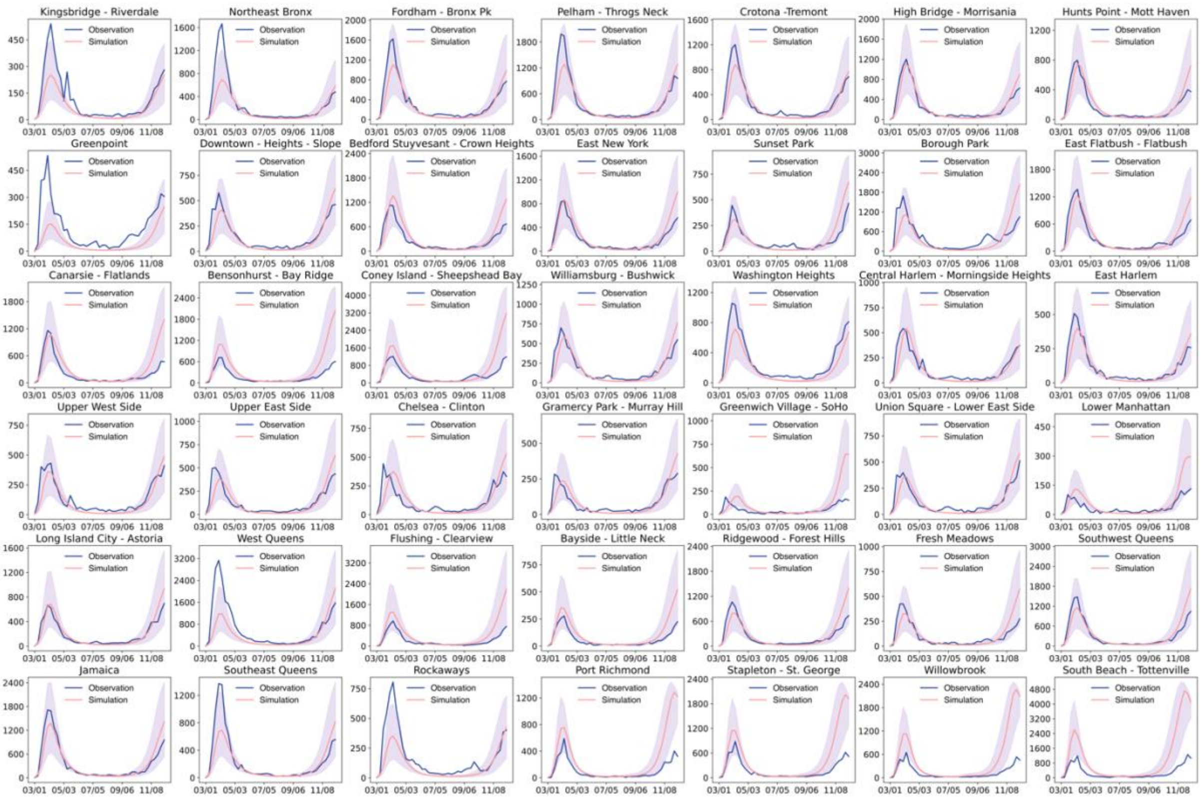


**Fig. S10. Model fitting in 42 NYC neighborhoods using the behavior-driven model without crowdedness and dwell time.** Model simulations using estimated posterior parameters (blue) are compared with the reported weekly cases in each neighborhood. Red lines are the observed weekly cases in NYC neighborhoods. The blue shaded areas show 95% CIs, obtained from 500 independent simulations, without adding the Gaussian observation error. The solid blue lines show the medians of 500 simulations.

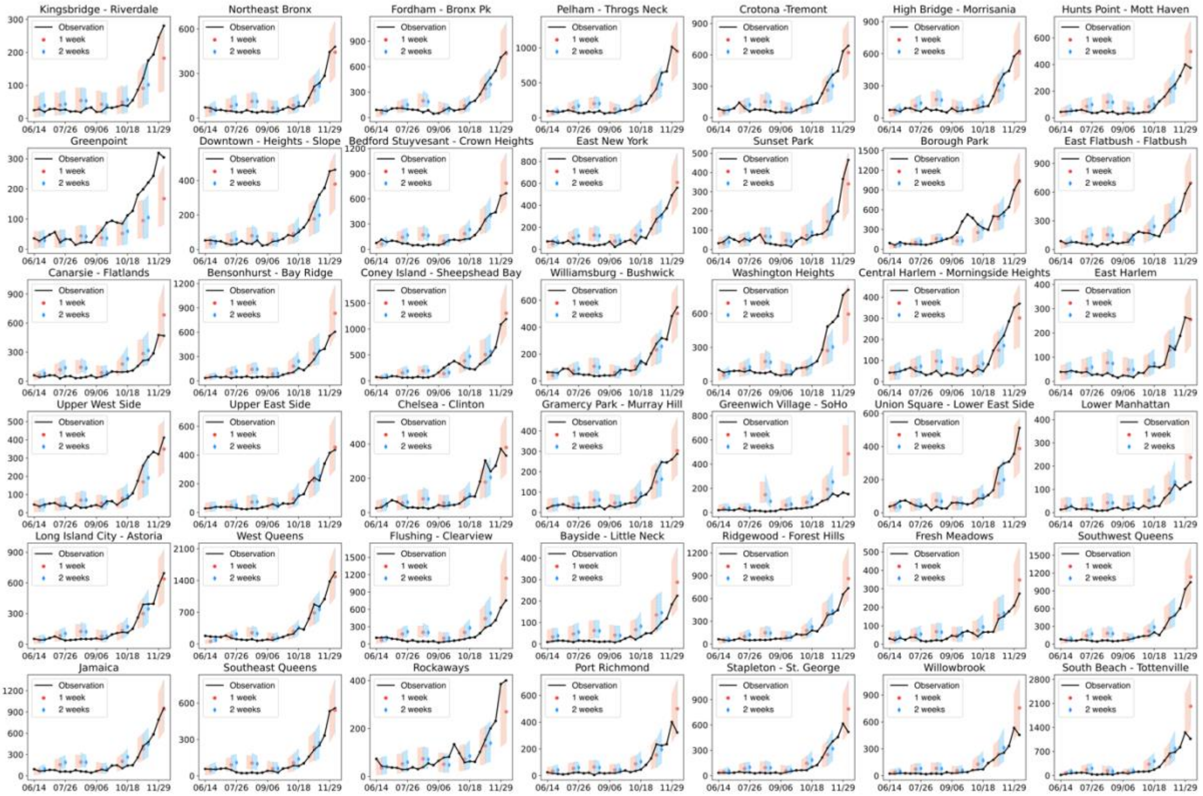


**Fig. S11. One-week and two-week ahead forecasts using the behavior-driven epidemic model.** We visualize forecasts generated at different weeks. The red and blue shaded areas show the one-week and two-week forecasts, respectively. Shaded areas are 95% CIs obtained from 500 ensemble members.

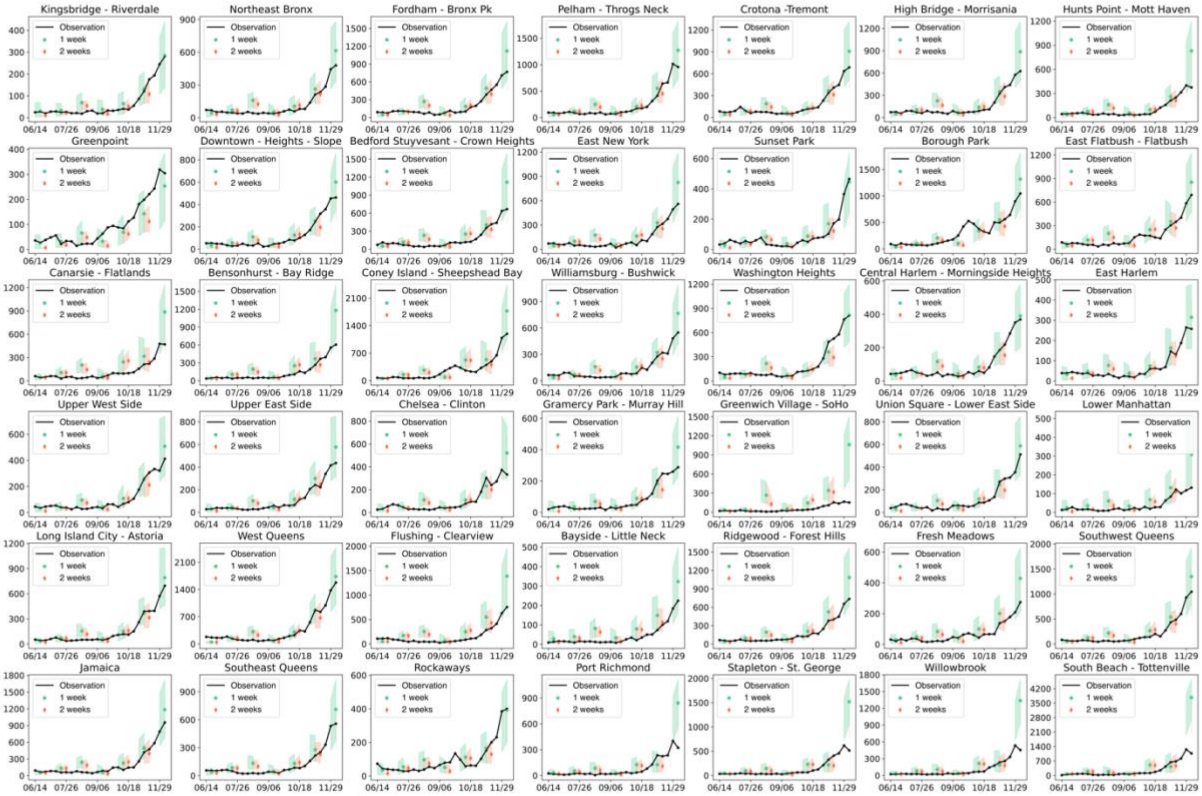


**Fig. S12. One-week and two-week ahead forecasts using the baseline model B1 (a metapopulation model without place category-specific mobility but with seasonal forcing).** We visualize forecasts generated at different weeks. The green and red shaded areas show the one-week and two-week forecasts, respectively. Shaded areas are 95% CIs obtained from 500 ensemble members.

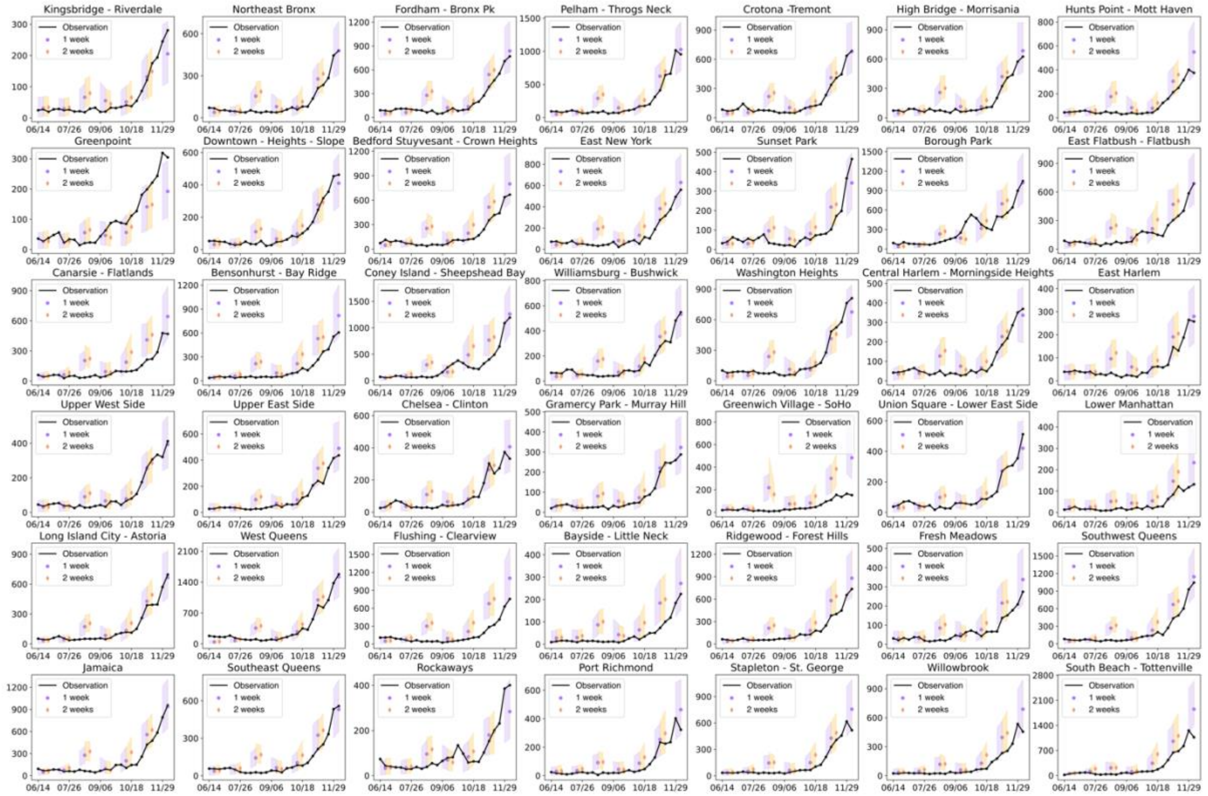


**Fig. S13. One-week and two-week ahead forecasts using the baseline model B2 (the behavior-driven model without seasonal forcing).** We visualize forecasts generated at different weeks. The purple and red shaded areas show the one-week and two-week forecasts, respectively. Shaded areas are 95% CIs obtained from 500 ensemble members.

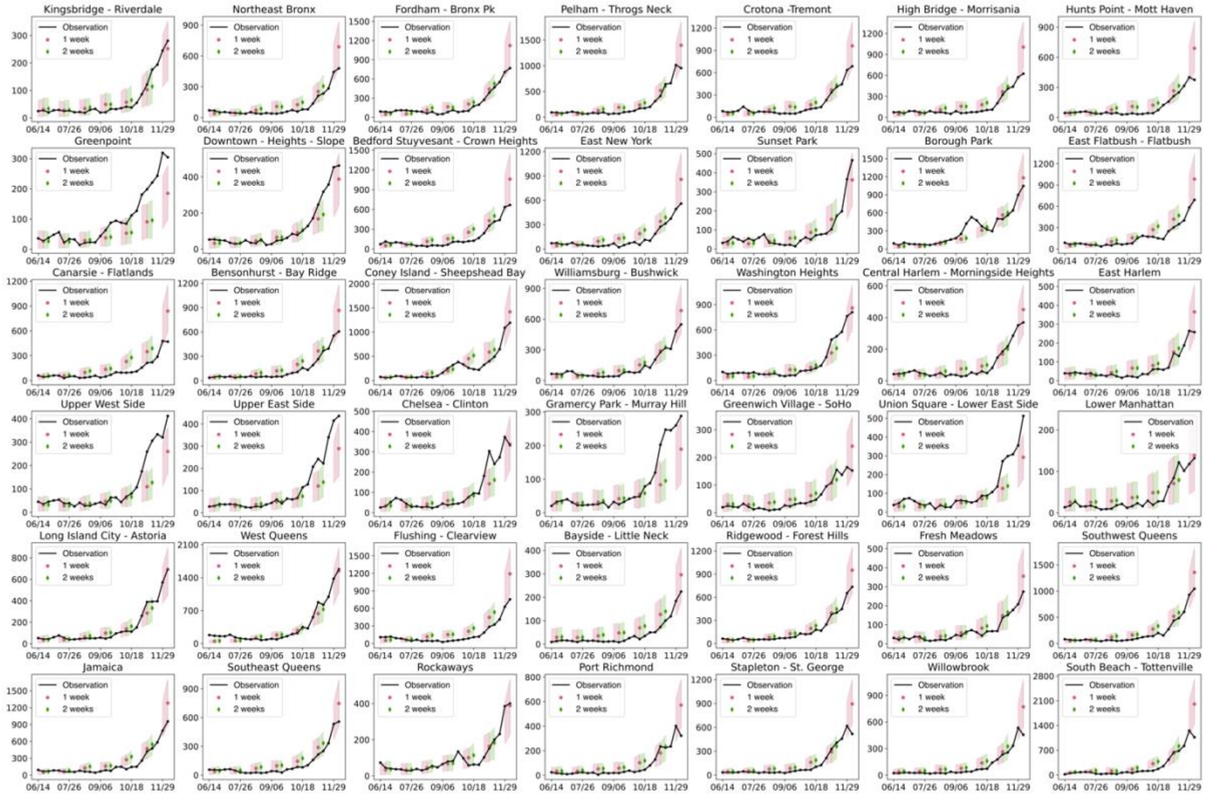


**Fig. S14. One-week and two-week ahead forecasts using the baseline model B3 (the behavior driven model with seasonal forcing but static mobility matrices, crowdedness, and dwell time).** We visualize forecasts generated at different weeks. The red and green shaded areas show the one-week and two-week forecasts, respectively. Shaded areas are 95% CIs obtained from 500 ensemble members.

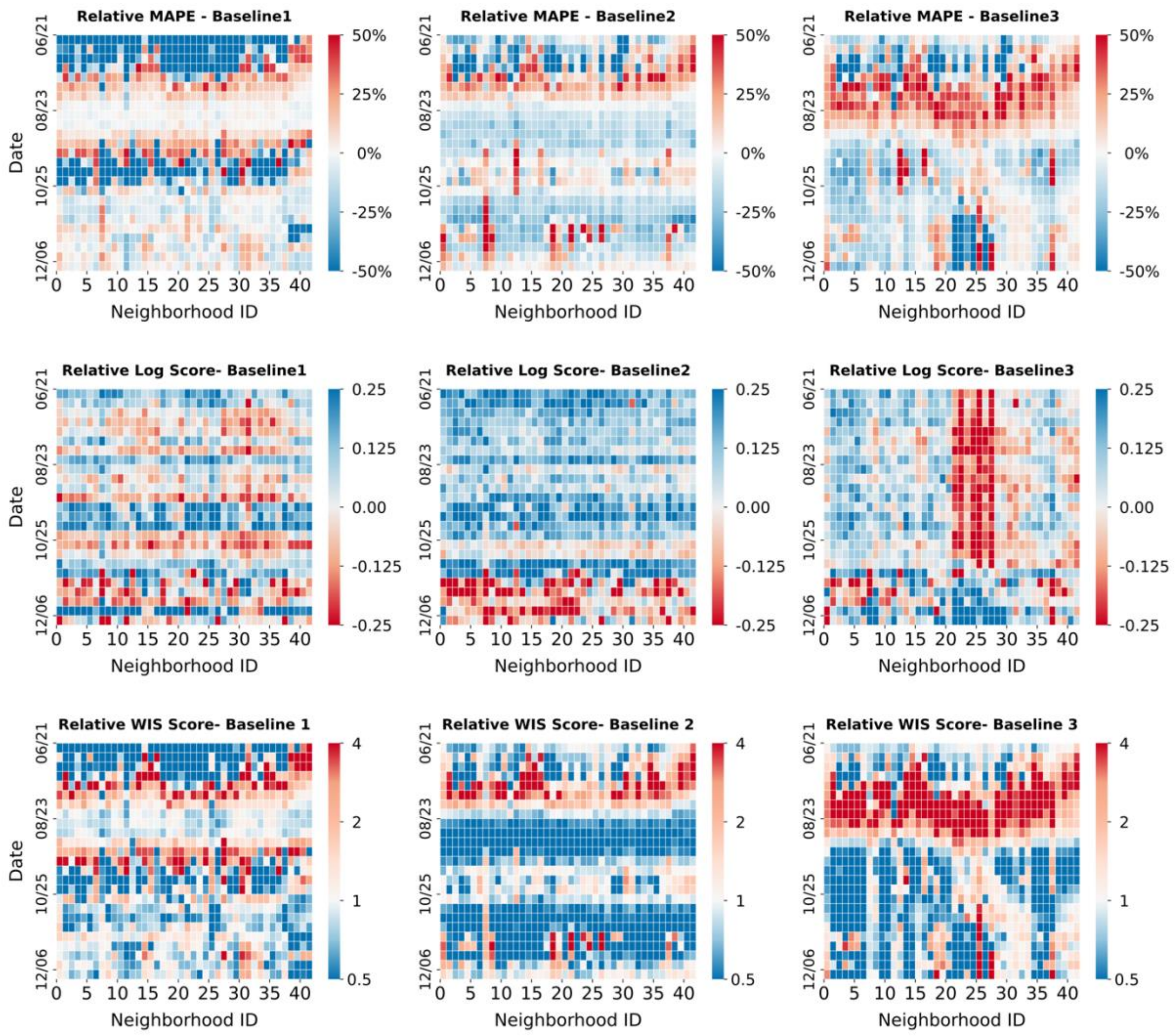


**Fig. S15. Comparison of two-week ahead forecasts.** The behavior-driven forecasts are compared with three baseline models: (B1), a metapopulation model without place category-specific mobility but with seasonal forcing; (B2), the behavior-driven model without seasonal forcing; (B3), the behavior driven model with seasonal forcing but static mobility matrices, crowdedness, and dwell time. We present (top row) the relative mean absolute percentage error (MAPE) (MAPEs of the behavior-driven model minus those of the baselines, with blue indicating better forecasts), (middle row) relative log score (log scores of the behavior-driven model minus those of the baselines, with blue indicating better forecasts), and (bottom row) relative weighted interval score (WIS) (the *ratio* of the WIS scores of the behavior-driven model to those of the baselines, with blue indicating better forecasts) for all 42 neighborhoods from June 8, 2020 to December 13, 2020.

**Table S1. Classification of POIs using the NAICS codes.**

| Place category | NAICS code and description | |
|---|---|---|
| Restaurants & bars | 7223 | Special Food Services |
| | 7224 | Drinking Places (Alcoholic Beverages) |
| | 7225 | Restaurants and Other Eating Places |
| Arts & entertainment | 7111 | Performing Arts Companies |
| | 7113 | Promoters of Performing Arts, Sports, and Similar Events |
| | 7131 | Amusement Parks and Arcades |
| | 7139 | Other Amusement and Recreation Industries |
| | 7112 | Spectator Sports |
| | 7121 | Museums, Historical Sites, and Similar Institutions |
| | 7132 | Gambling Industries |
| Educational settings | 6111 | Elementary and Secondary Schools |
| | 6112 | Junior Colleges |
| | 6113 | Colleges, Universities, and Professional Schools |
| | 6115 | Technical and Trade Schools |
| | 6116 | Other Schools and Instruction |
| Retail | 4411 | Automobile Dealers |
| | 4412 | Other Motor Vehicle Dealers |
| | 4413 | Automotive Parts, Accessories, and Tire Stores |
| | 4421 | Furniture Stores |
| | 4422 | Home Furnishings Stores |
| | 4431 | Electronics and Appliance Stores |
| | 4441 | Building Material and Supplies Dealers |
| | 4442 | Lawn and Garden Equipment and Supplies Stores |
| | 4451 | Grocery Stores |
| | 4452 | Specialty Food Stores |
| | 4453 | Beer, Wine, and Liquor Stores |
| | 4461 | Health and Personal Care Stores |
| | 4471 | Gasoline Stations |
| | 4481 | Clothing Stores |
| | 4482 | Shoe Stores |
| | 4483 | Jewelry, Luggage, and Leather Goods Stores |
| | 4511 | Sporting Goods, Hobby, and Musical Instrument Stores |
| | 4512 | Book Stores and News Dealers |
| | 4522 | Department Stores |
| | 4523 | General Merchandise Stores, including Warehouse Clubs and Supercenters |
| | 4531 | Florists |
| | 4532 | Office Supplies, Stationery, and Gift Stores |
| | 4533 | Used Merchandise Stores |
| | 4539 | Other Miscellaneous Store Retailers |
| Other places | Other NAICS codes | |
